# Deep learning on CT scans to predict checkpoint inhibitor treatment outcomes in advanced melanoma

**DOI:** 10.1101/2023.07.25.23293133

**Authors:** L.S. Ter Maat, R.A.J. De Mooij, I.A.J. Van Duin, J.J.C. Verhoeff, S.G. Elias, T. Leiner, W.A.C. van Amsterdam, M.F. Troenokarso, E.R.A.N. Arntz, F.W.P.J. Van den Berkmortel, M.J. Boers-Sonderen, M.F. Boomsma, A.J.M. Van den Eertwegh, J.W. De Groot, G.A.P. Hospers, D. Piersma, G. Vreugdenhil, H.M. Westgeest, E. Kapiteijn, G.A. De Wit, W.A.M. Blokx, P.J. Van Diest, P.A. De Jong, J.P.W. Pluim, K.P.M. Suijkerbuijk, M. Veta

**Author notes:** These authors contributed equally (K.P.M. Suijkerbuijk, M. Veta).

## Abstract

**Introduction:** Checkpoint inhibitor treatment has proven successful for advanced melanoma. However, a significant fraction of patients does not experience benefit from this treatment, that is also associated with potentially severe toxicity and high costs. Previous research has not yet resulted in adequate biomarkers that can predict treatment outcomes. The present work is the first to investigate the value of deep learning on computed tomography (CT) imaging of melanoma lesions for predicting checkpoint inhibitor treatment outcomes in advanced melanoma.

**Methods:** Adult patients that were treated with first line anti-PD1 ± anti-CTLA4 therapy for unresectable stage IIIC or stage IV melanoma were retrospectively identified from ten participating centers. Up to five representative lesions were segmented volumetrically on baseline CT; a deep learning model (DLM) was trained on the corresponding volumes to predict clinical benefit, defined as stable disease for a minimum of six months, or response at any time during follow-up. Optimal hyperparameters and model types (Densenet, Efficientnet, Squeeze-Excitation ResNet, ResNeXt) were iteratively explored. The DLM was compared to a model of previously identified clinical predictors (presence of liver and brain metastasis, level of lactate dehydrogenase, performance status and number of affected organs), and a combination model consisting of both clinical predictors and the DLM.

**Results:** A total of 730 eligible patients with 2722 lesions were included. Rate of clinical benefit was 59.6%. The selected deep learning model was a Squeeze-Excitation ResNet with random initialization, trained with the Adam optimizer. The DLM reached an area under the receiver operating characteristic (AUROC) of 0.607 [95% CI 0.565 – 0.648]. In comparison, a model of clinical predictors reached an AUROC of 0.635 [95% CI 0.592 – 0.678]. The combination model reached an AUROC of 0.635 [95% CI 0.595 – 0.676]. None of the differences in AUROC were statistically significant. The output of the DLM was significantly correlated with four of the five input variables of the clinical model.

**Discussion:** Although the DLM reached a statistically significant discriminative value, it was unable to improve over previously identified clinical predictors. The most likely cause is that the DLM learns to detect a lesion’s size and organ location, which is information that is already present in the clinical model. Given the substantial sample size and extensive hyperparameter optimization, this indicates that the predictive value of CT imaging of lesions for checkpoint inhibitor response in melanoma is likely limited. The present work shows that the assessment over known clinical predictors is an essential step for imaging-based prediction and brings important nuance to the almost exclusively positive findings in this field.

## Introduction

**Checkpoint inhibitors have revolutionized the treatment of advanced melanoma**. The real-world 1-year overall survival of patients treated with anti-PD1 therapy is 67% [1], which is in stark contrast to the 1-year overall survival of 25% in phase II trials up to 2007 [2].

**However, still a significant fraction of patients does not respond to this treatment, that is also associated with potentially severe toxicity and high costs**. Approximately 40-50% of patients experience disease progression despite treatment, and subsequently derive little benefit in terms of survival [1,3]. Furthermore, checkpoint inhibition treatment is expensive, with estimates of additional costs of up to 81,000 US dollars per quality adjusted life year [4,5]. Lastly, severe and partly irreversible toxicity occurs in as much as 60% of patients treated with anti-PD1 + anti-CTLA4 combination therapy [6].

**Therefore, accurate prediction at baseline of treatment outcomes is necessary**. If non-responders can be identified with high certainty before start of treatment, alternative therapies can be started without delay in these patients. Furthermore, needless costs and toxic effects can be prevented.

**However, current biomarkers are not accurate enough to guide treatment decisions**. Previous research has identified several significant predictors of treatment outcomes, such as levels of lactate dehydrogenase, presence of liver and brain metastases, performance status and level of tumoral PD-L1 expression [7,8]. These biomarkers, however, have not reached the degree of accuracy that is necessary to adequately guide treatment decisions. Patients without PD-L1 expression, for instance, may still respond to therapy, even though this protein is the very target of anti-PD1 therapy [8]. This underlines the need for further research into accurate predictive biomarkers.

**CT imaging of tumor lesions may be used as a biomarker in two ways: through handcrafted radiomics and through deep learning**. In a handcrafted radiomics approach, predefined features that reflect shape and texture are calculated on a volume of interest. These features are subsequently used to train a model that can classify the lesion as, for instance, having a certain mutation or responding to a treatment [9]. In contrast, a deep learning approach skips the step of extracting manually predefined features and trains a model directly on the raw image as an input [10]. This approach has the advantage that it is not limited by the chosen features in what it can learn; instead, relevant features are learned during training in such a way that the predictive performance of the model is optimized. A potential downside is that usually a larger dataset is needed for adequate performance compared with a handcrafted radiomics approach. For both methods, the underlying hypothesis is that features visible on imaging reflect the tumor’s phenotype and may therefore also correlate to clinically relevant characteristics and biological behavior of the tumor.

**Thus far, deep learning on CT imaging of lesions has not been investigated for predicting checkpoint inhibitor treatment outcomes in melanoma patients**. Previous studies have investigated the use of deep learning on CT imaging for this purpose in other malignancies, namely non-small cell lung carcinoma (NSCLC) [11–14] and urothelial carcinoma [15,16], with positive findings. For melanoma, only handcrafted radiomics have been investigated thus far [17–20]. Initial findings by other smaller, single-center studies were promising, but our recent study of 620 patients from nine different centers showed different results: although the radiomics model had some value in predicting ICI treatment outcomes, it did not outperform a model based on clinical characteristics [20]. Deep learning may improve the performance over handcrafted radiomics as it is not limited by the choice of predefined features. This hypothesis remains to be experimentally verified, as studies comparing handcrafted radiomics to deep learning for other tasks show conflicting results [21–24].

**The aim of this work was to determine the added value of deep learning on baseline CT imaging of lesions over clinical predictors for predicting first-line checkpoint inhibitor treatment outcomes in patients with advanced cutaneous melanoma**. We have collected and curated a multi-center dataset of baseline CT imaging of these patients specifically for this purpose. With a sample size of 716 patients and 2722 lesions, this dataset is currently the largest of its kind in melanoma, and among the largest in all cancer types for imaging-based prediction of checkpoint inhibitor treatment outcomes [25].

## Methods

### Patient selection

Eligible patients were retrospectively identified from 10 participating centers (Amphia Ziekenhuis, Isala Zwolle, LUMC, Máxima MC, Medisch Spectrum Twente, Radboudumc, UMC Groningen, UMC Utrecht, Amsterdam UMC, Zuyderland MC) using prospectively collected high-quality registry data. With the exception of the UMC Groningen, this is the same population as in a previous work, which investigated handcrafted radiomics for the same purpose [20]. Patients were eligible if they were (i) treated for unresectable stage IIIC or IV cutaneous melanoma (ii) using first-line anti-PD1 ± anti-CTLA4 checkpoint inhibition (iii) on or after 1-1-2016 and (iv) were over 18 years of age at the start of treatment. Exclusion criteria were (i) unavailability of baseline contrast-enhanced CT imaging and (ii) absence of eligible lesions on CT.

### ROI selection and preprocessing

Up to five lesions per patient were selected and manually segmented by authors LSM and IAJD under supervision of board-certified radiologists with 17 and 18 years of experience (PJ and TL, respectively). First, the five largest lesions were segmented with a maximum of two lesions per organ. Then, if fewer than five lesions had been segmented but more lesions remained, the largest remaining lesions were segmented up to a maximum of five. For example: in a patient with five large lung lesions and one small liver lesion, the two largest lung lesions and single liver lesion are segmented first. Then, the two largest remaining lung lesions are segmented, resulting in a total of five segmented lesions. Regions of interest (ROI) were extracted as cubes centered on the centroid of the segmentation. During training and validation steps, the data was augmented through random rotation around all spatial axes and addition of Gaussian noise.

### Outcome definition

The primary outcome was clinical benefit, defined as a best overall response of ‘stable disease’ for a minimum of six months, or ‘partial response’ or ‘complete response’, as determined by the treating physician in line with RECIST 1.1 criteria [26]. The secondary outcome was objective response, defined as a best overall response of ‘partial response’ or ‘complete response’. In addition, lesion outcomes were determined based on maximum diameter measurements at baseline, 3, 6 and 9 months. If the maximum diameter at the last available measurement exceeded 120% of the original maximum diameter, the lesion was labeled as ‘no benefit’, and otherwise as ‘benefit’. Similarly, lesions were labeled as ‘response’ or ‘no response’ using a 70% cut-off. Both cut-offs were chosen in line with the RECIST 1.1 criteria for determining patient response.

### Model selection and hyperparameter selection

To arrive at a well-optimized model, a range of options for certain design choices (so-called ‘hyperparameters’) were systematically explored. These hyperparameters included, among others, model architecture, learning rate and choice of optimizer. For model architectures, considered options were ResNet [27], Squeeze-Excitation ResNet [28], EfficientNet [29] and ResNeXt [30]. A full list of all hyperparameters along with possible values is supplied in Supplementary Table 1. To efficiently explore the vast space of possible hyperparameter combinations, an iterative process was used. In every iteration, a small number of hyperparameters were investigated using a random search strategy and a randomly chosen fixed train-validation split. The values with the highest validation area under the receiver operating characteristic (AUROC) for predicting patient level outcomes were subsequently fixed. This process was repeated until optimal values were selected for all hyperparameters. An iteration was continued for a maximum of 100 epochs, with early stopping after 10 epochs of no improvement of the patient level area under the curve (AUC) on the validation set.

### Model training and evaluation

The selected configuration of model and hyperparameters was evaluated using a nested cross validation. The inner loop was conducted in a 5-fold cross validation. In every fold, 80% of the patients made up the training data; the remaining 20% was used as a validation set for monitoring training and early stopping. Repeating this process in all five folds resulted in five trained models, which were used in an ensemble: a combined model that averages the predictions of the five models per lesion. The outer loop was conducted in a leave-one-center-out manner and was used to evaluate the performance of the ensemble on an independent test set.

During training, the model was optimized to predict the lesion level outcome based on the ROI of the corresponding lesion. During inference, these lesion level predictions were aggregated to a patient level by taking the minimum, mean or maximum of all predictions for a single patient. The choice for minimum, mean or maximum was also considered a hyperparameter. For predicting patient clinical benefit, lesion benefit was used as the lesion level label; for predicting patient objective response, lesion response was used as the lesion level label. Lesions with unavailable lesion level outcomes could not be used during training; these lesions were used during inference, however, as only patient level outcomes were necessary at this stage.

The model was compared to a previously published clinical model [20] and a combination model of both the deep learning model and clinical model. The clinical model was a logistic regression based on four variables which were previously shown to be significant predictors of checkpoint inhibitor treatment outcomes in patients with advanced melanoma [1,7,31]. These predictors were presence of (i) liver and (ii) (a)symptomatic brain metastases, (iii) Eastern Cooperative Oncology Group (ECOG) performance status and (iv) levels of lactate dehydrogenase (LDH). Further details of the clinical and combination model are available in the Supplementary Methods.

### Statistical analysis

Model calibration was assessed using calibration curves and Hosmer-Lemeshow test. Model discrimination was assessed using the receiver-operator characteristics (ROC) curve and corresponding AUC; 95% confidence intervals were calculated using the cvAUC R package [32]. Methods for comparing cross validated AUCs are described in the Supplementary Methods. The learned representation of the deep learning model was visualized using a two-dimensional t-distributed stochastic neighbor embedding (t-SNE).

### Adherence to quality standards

After review by the Medical Ethics Committee, this study was deemed not subject to the Medical Research Involving Human Subjects Act in accordance with Dutch regulations. Informed consent was waived.

## Results

### Patient characteristics

Out of 1347 eligible patients, 617 patients were excluded, resulting in 730 included patients with 2722 lesions; most exclusions were due to the availability of only a low-dose CT from a combined FDG-PET scan, instead of a diagnostic CT scan (Figure 1). 59.6% of 730 patients had clinical benefit (435 patients); the objective response rate was 51.1% (373 patients). Outcomes for individual lesions were available in 2128 lesions (78.2%); 21.8% of lesion outcomes were unavailable due to local regulations in one hospital (12.0%, 327 lesions), due to death or clinical progression before the first follow-up moment (7.4%, 202 lesions), due to the lesion falling outside the field-of-view of the scan (0.8%, 21 lesions) or due to technical problems (1.6%, 44 lesions); availability of lesion outcomes at 3, 6 and 9 months is shown in Supplementary Table 2. Among lesions with available outcomes, the rate of benefit was 79.7%; the lesion response rate was 55.2%. Characteristics of included and excluded patients are displayed in Table 1 and Supplementary Table 3. Included patients had on average more advanced disease than excluded patients. Acquisition parameters and patient characteristics per center and subgroup are shown in Supplementary Tables 4-6.

**Figure 1.**
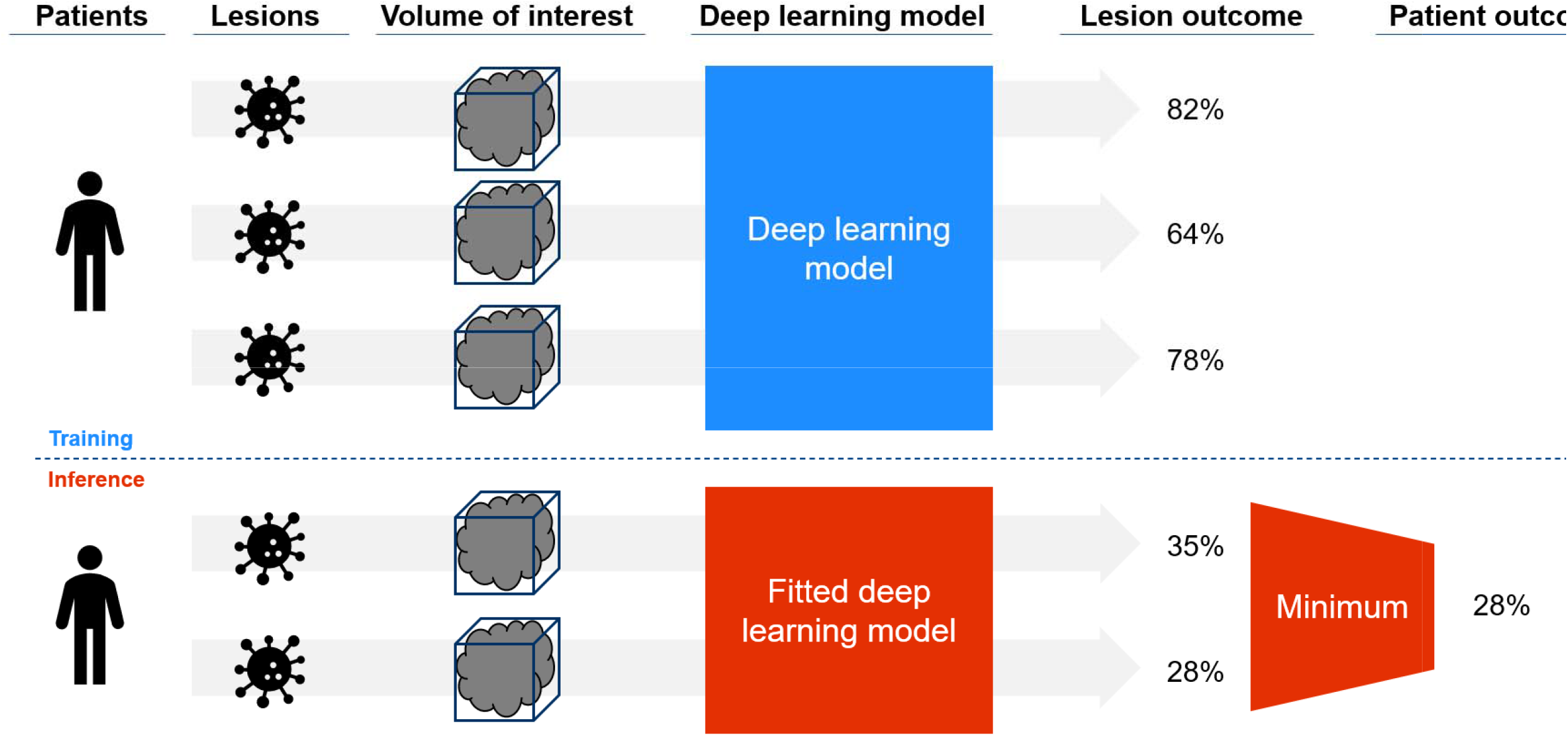
Graphical overview of the proposed method. From left to right: for every eligible patient, up to five representative lesions are selected. A 3D volume of interest on the pretreatment CT scan is used as input for the deep learning m During training (above the dotted line), the deep learning model is optimized to predict the probability of benefit from checkpoint inhibition for every individual lesion. During inference (belo dotted line), the fitted deep learning model is used to make lesion level predictions. These lesion level predictions are subsequently aggregated to a patient level prediction. Several options explored for how to aggregate lesion level predictions, namely by taking the maximum, mean or minimum of predictions. After hyperparameter tuning, the ‘minimum’ function was selected.

**Figure 2.**
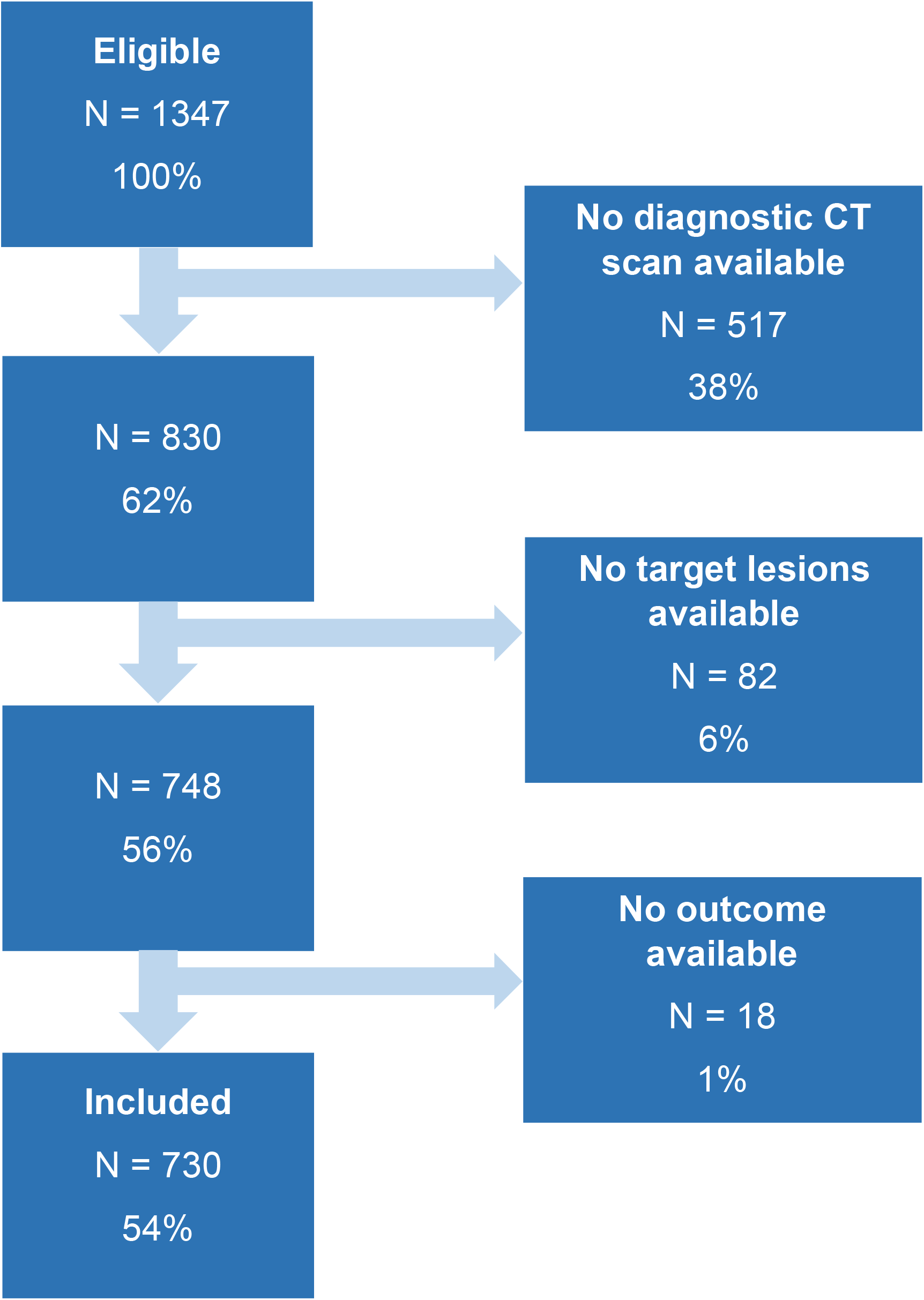
Flowchart of the inclusion process.

**Table 1.** Characteristics of included patients.

| n |  | 730 |
| --- | --- | --- |
| Age, median [Q1,Q3] |  | 68.0 [58.0,75.0] |
| Sex, n (%) | Female | 285 (39.0) |
|  | Male | 445 (61.0) |
| Therapy, n (%) | Anti-PD1 | 458 (62.7) |
|  | Ipilimumab & Nivolumab | 272 (37.3) |
| Stage, n (%) | IIIC | 28 (3.8) |
|  | IV M1a | 56 (7.7) |
|  | IV M1b | 114 (15.6) |
|  | IV M1c | 344 (47.1) |
|  | IV M1d | 182 (24.9) |
|  | Missing | 6 (0.8) |
| ECOG performance status, n (%) | 0 | 356 (48.8) |
|  | 1 | 271 (37.1) |
|  | 2-4 | 73 (10.0) |
|  | Missing | 30 (4.1) |
| Brain metastases, n (%) | absent | 497 (68.1) |
|  | asymptomatic | 94 (12.9) |
|  | symptomatic | 88 (12.1) |
|  | missing | 51 (7.0) |
| Liver metastases, n (%) | absent | 471 (64.5) |
|  | present | 224 (30.7) |
|  | missing | 35 (4.8) |
| LDH, n (%) | normal | 459 (62.9) |
|  | 1-2x ULN | 199 (27.3) |
|  | >2x ULN | 62 (8.5) |
|  | missing | 10 (1.4) |
| Number of affected organs, n (%) | <3 | 432 (59.2) |
|  | ≥3 | 298 (40.8) |
| Best overall response, n (%) | Complete response | 94 (12.9) |
|  | Partial response | 279 (38.2) |
|  | Stable disease | 115 (15.8) |
|  | Progressive disease | 237 (32.5) |
|  | Death | 5 (0.7) |
| Clinical benefit, n (%) | benefit | 435 (59.6) |
|  | no benefit | 295 (40.4) |
| Objective response, n (%) | response | 373 (51.1) |
|  | no response | 357 (48.9) |

**Abbreviations**
ECOG=Eastern Cooperative Oncology Group LDH=lactate dehydrogenase, ULN=upper limit of normal, defined as 250 IU/L

### Hyperparameter selection

Ten iterations of preliminary experiments were performed; the results are available online through Supplementary Table 6. Based on these experiments, the model architecture was set to the Squeeze-Excitation ResNet50 [28] model with 3-dimensional input and random initial parameters; the function for aggregating predictions of all lesions belonging to one patient was selected to be ‘minimum’. The Adam optimizer was used with a cosine annealing learning rate scheduler. Other hyperparameters are listed in Supplementary Table 1.

### Treatment outcome prediction

The deep learning model achieved a leave-one-center-out cross-validated AUROC of 0.607 [95% CI 0.565-0.648] for predicting clinical benefit. In comparison, the clinical model achieved an AUROC of 0.635 [95% CI 0.592-0.678], and the combination model an AUROC of 0.635 [95% CI 0.595-0.676]. Differences in AUROC between the clinical and combination model were not statistically significant (Supplementary Figures 3). There was no evidence of poor fit in the three models (Hosmer-Lemeshow p > 0.113). The 95% interval of predicted probabilities was 0.51-0.63 for the deep learning model, 0.28-0.77 for the clinical model and 0.49-0.72 for the combination model. Results were similar for prediction of objective response (Supplementary Figure 4 and 5), and in treatment subgroups (Supplementary Figure 6 – 9).

### Interpretability analysis

Figure 5 shows the t-SNE embedding of the final layer of one of the fitted models (outer fold ‘Amsterdam UMC’, inner fold 3). The t-SNE analysis shows that the deep learning model learns to detect a lesion’s organ location (Figure 5A). Especially for liver and lung lesions, the predicted probability of lesion benefit is lower and higher, respectively (Figure 5D). However, there is a large overlap between benefitting and non-benefitting lesions (Figure 5B). Supplementary Figures 11 and 12 show the same analysis for different outer and inner folds. In line with these findings, Figure 6 shows that the patient level predictions of the deep learning model are significantly correlated with four out of five of the clinical predictors (Kruskal-Wallis p < 0.020). Furthermore, lesion level predictions are weakly but significantly correlated with lesion volume (r = −0.28, p < 0.0001).

**Figure 3.**
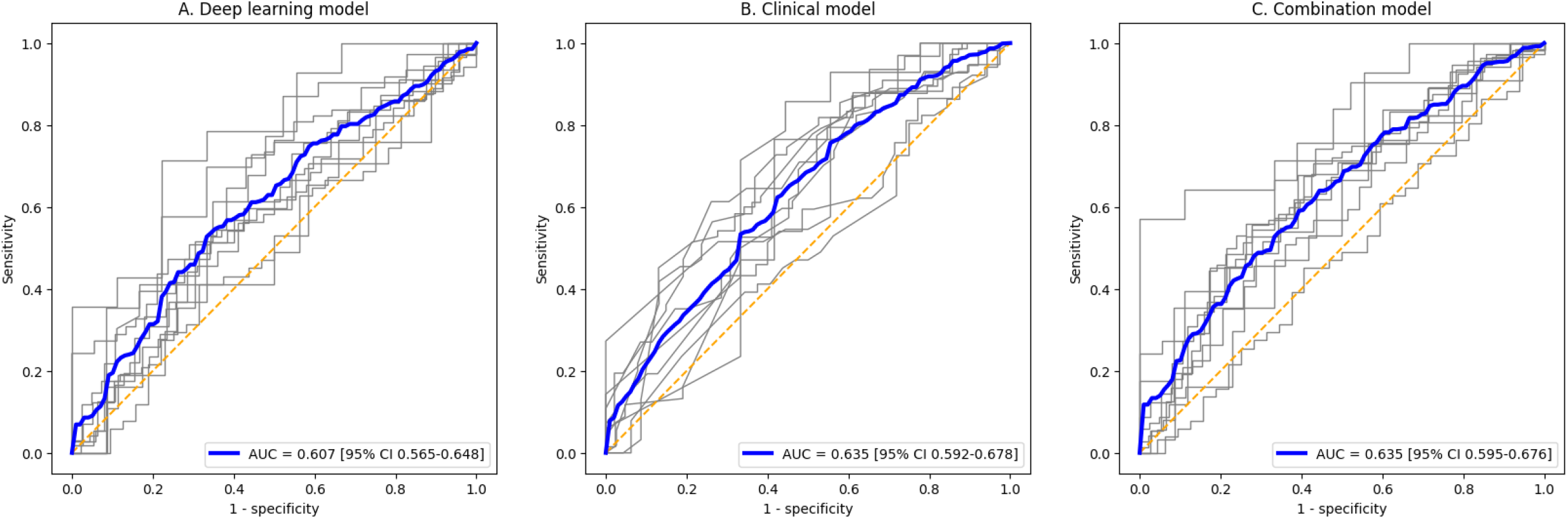
Receiver operator characteristic curves for predicting clinical benefit. Receiver operator characteristic (ROC) curves for (A) the deep learning model, (B) the baseline clinical model and (C) the combination model for predicting clinical benefit on a patient level. Curves of the individual folds/validation centers are shown in gray; the average ROC curve is shown in blue. Corresponding areas under the curve (AUC) are supplied in the legend. The orange line corresponds to the line of random performance.

**Figure 4.**
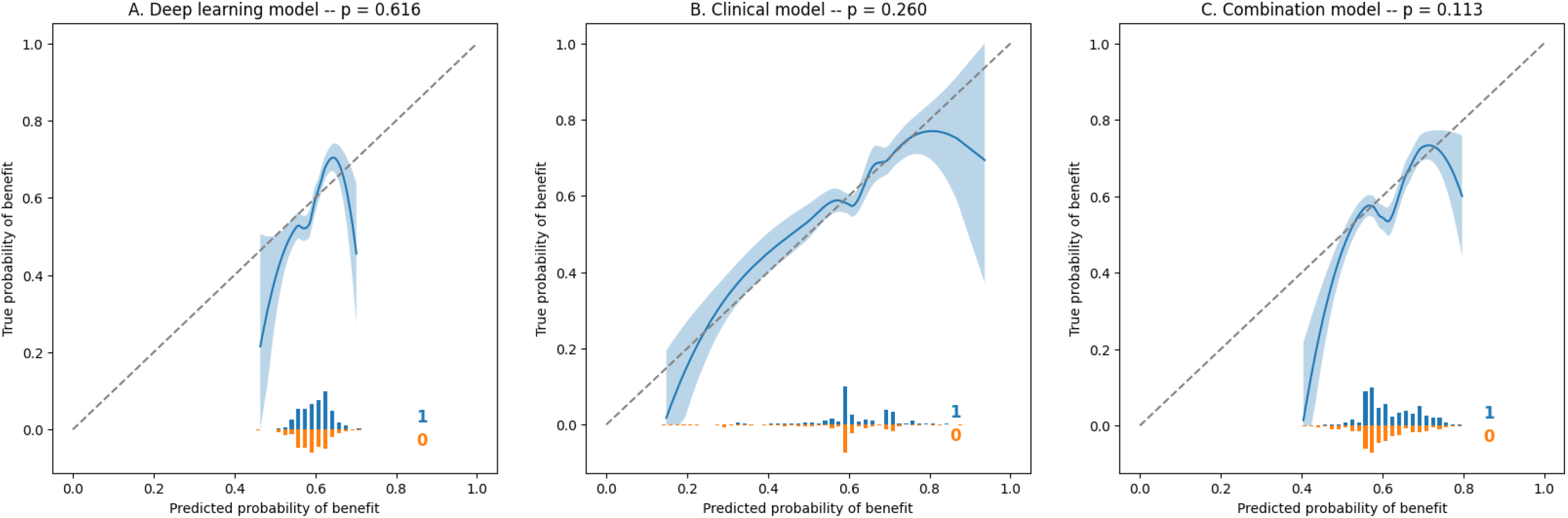
Calibration curves for predicting clinical benefit. Locally estimated scatterplot smoothing (LOESS) fitted calibration curves with corresponding 95% confidence interval for (A) the deep learning model, (B) the clinical model and (C) the combination model for predicting clinical benefit on a patient level. The dashed line indicates the line of perfect calibration. Histograms of individual predictions, split for patients with (blue) and without (orange) benefit, are shown below the curves. The p-value for the Hosmer-Lemeshow test for goodness of fit is shown in the plot title.

**Figure 5.**
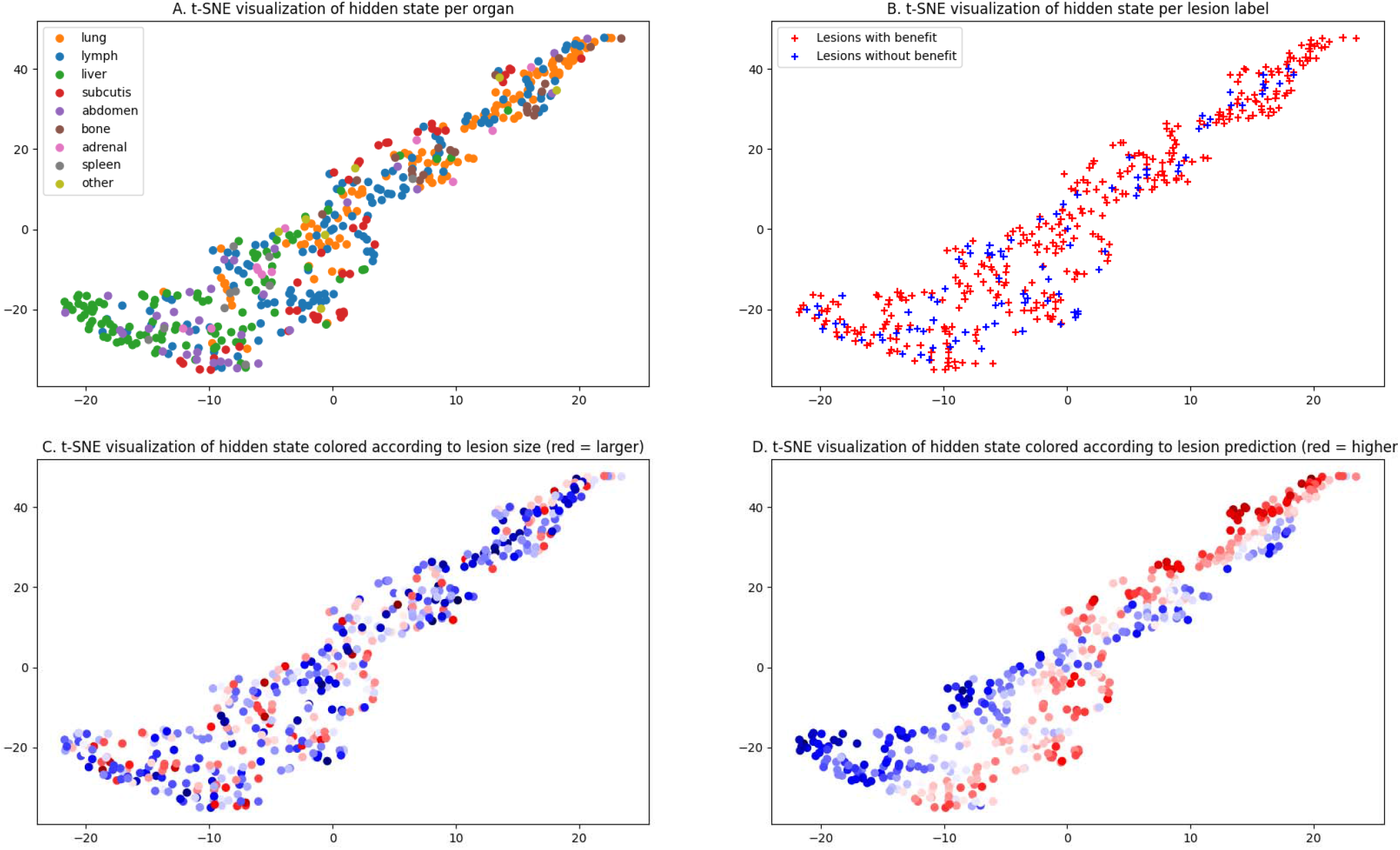
t-SNE analysis on lesion level of the representation learned by the deep-learning model for predicting clinical benefit (outer fold ‘Amsterdam UMC’, inner fold 3) Based on the training data, the deep learning model learns to map every lesion to a point in space where, intuitively, similar lesions are closer together. This mapping is visualized in this figure in 2D using t-SNE. Every point corresponds to a single lesion. Relative distance indicates how similar lesions are according to the model; absolute location is not informative in this figure. Lesions are colored in the four different plots to show how the information learned by the model corresponds with information about the lesion. (A) Lesions located in different organs are clustered together, indicating that the deep learning model detects the lesion’s location. (B) There is no clear separation of lesions with and without benefit, indicating that the model cannot accurately discriminate between lesion treatment outcomes. (C) Although some clusters of large and small lesions can be seen, lesion size appears to be less determining for the model’s output than location. (D) Overall, predicted probability of benefit is lower in lesions marked as liver lesions in Figure 4A, and higher in lung lesions.

**Figure 6.**
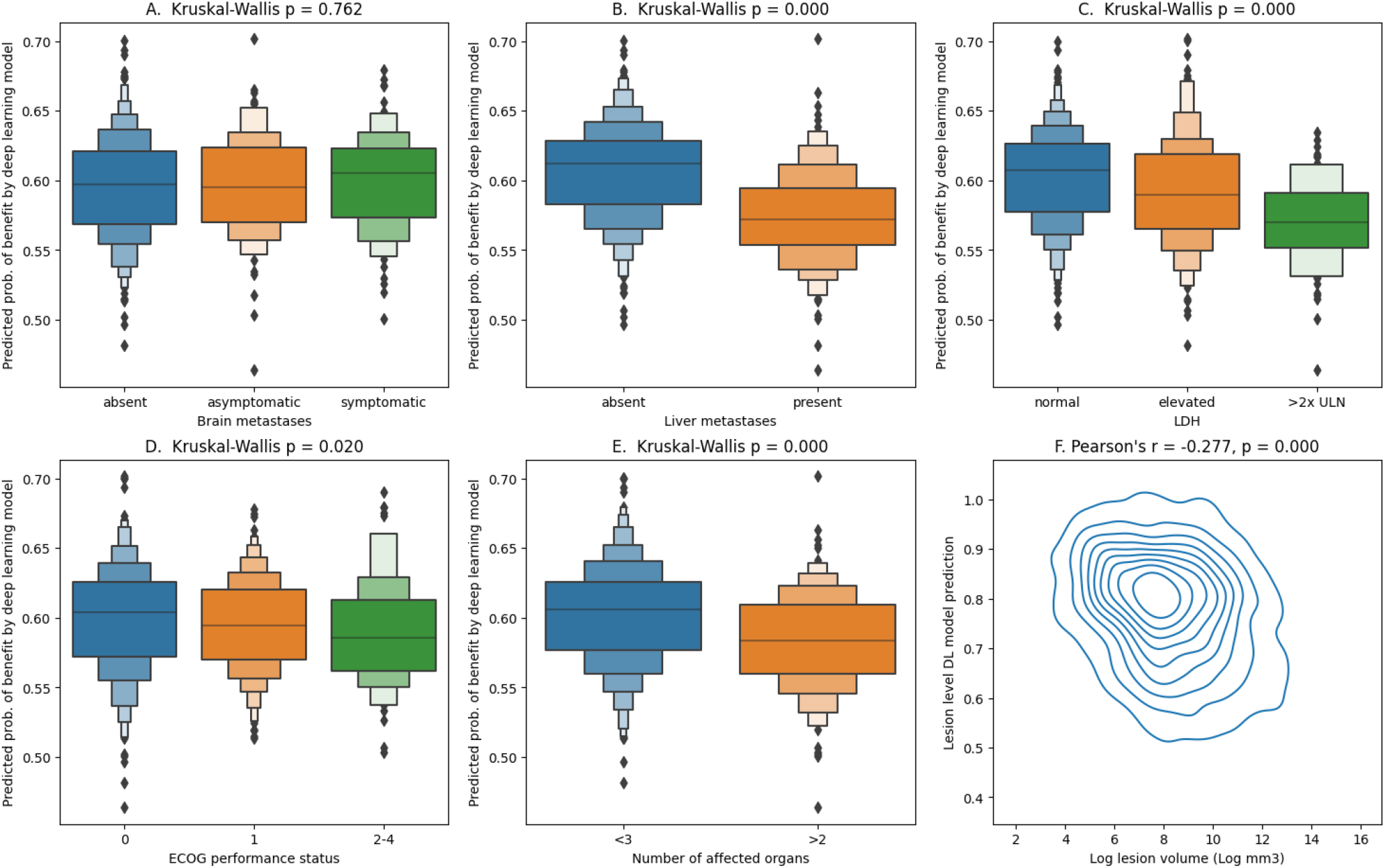
Patient level predictions of the deep learning model for probability of clinical benefit, compared across clinical variables. (A-E) Boxenplots of known clinical predictors with the output of the deep learning model for predicting clinical benefit per patient. P-values of the Kruskal-Wallis test for difference in distribution are given in the plot titles. (A) The output of the deep learning model is not significantly different for patients with or without brain metastases. (B-E) The output of the deep learning model is significantly different for patients with and without liver metastases (B), with varying levels of LDH (C), different ECOG performance status (D) and with less than 3 and 3 or more affected organs (E). (F) Kernel-density estimate plot of log-transformed lesion volume versus the lesion-level prediction of the deep learning model. The output of the deep learning model per lesion is significantly lower in larger lesions.

## Discussion

**A deep learning model on CT imaging of lesions had a significant but clinically limited predictive value for predicting response to checkpoint inhibitors in patients with advanced melanoma**. Despite the substantial dataset size and extensive hyperparameter tuning, the achieved level of discrimination was limited. This result, combined with earlier findings on handcrafted radiomics, indicates that CT imaging of melanoma lesions at baseline holds limited information about treatment outcomes. Other studies have demonstrated that using on-treatment scans yields substantially better predictive performance, but on-treatment prediction is clinically far less relevant: most toxicity occurs in the first three months [33], and conventional follow-up measurements can already accurately predict long-term outcomes [34].

**Addition of this deep learning model to clinical predictors did not improve predictive value**. The difference in discrimination between both models was marginal. This was despite the large sample size and the cross-validation setup, which leverages every patient for independent validation. Furthermore, the range of predicted probabilities was wider for the clinical model.

**This overlap in predictive value is likely to stem from the fact that the deep learning model learns information which is already encoded in the clinical model**. The most plausible explanation is that the model encodes a lesion’s size and organ location, which may subsequently be correlated with stage and tumor load and therefore LDH, ECOG performance status and number of affected organs. This is in line with our earlier findings using a handcrafted radiomics approach [20].

**The present work has important implications for future research**. First, the overlap in predictive information between the clinical and deep learning model shows that it is essential to assess the added value of an imaging-based model over known predictors. In practice, however, this is rarely done [25]. Second, the present work suggests that previous results on imaging-based prediction of checkpoint inhibitor outcomes may be overoptimistic. Published results are almost exclusively positive, but numerous concerns exist regarding study size and quality [25]. The fact that these positive results are not confirmed in a large, multicenter dataset curated specifically for this purpose nuances this optimism.

**The strengths of the present work are the large sample size and multicenter design**. The training of deep learning models requires a substantial dataset size due to the large number of trainable parameters. To our knowledge, we have collected the largest dataset to date. Furthermore, the multicenter design allows for the evaluation of the generalizability of the model to new centers, which was a limitation of most previous studies. This, in combination with the cross-validation setup, adds significantly to the strength of the presented analysis.

**This study has two potential limitations**. First, a large group of patients was excluded due to unavailability of a contrast-enhanced baseline CT scan. Our hypothesis for the small difference in disease stage between in- and excluded patients is that patients with more advanced disease are more likely to present to medical oncology directly, instead of being referred after an FDG-PET CT scan has been performed. The risk of selection bias is limited however, as absolute differences in characteristics between in- and excluded patients are small. Second, performance of the deep learning model could in theory improve with the inclusion of more than five lesions per patient. However, we believe this is unlikely to change the conclusion, as a sensitivity analysis in a subset of the data did not show a difference in performance when more lesions were included. Furthermore, more than half of patients have at most five lesions.

**In conclusion, a deep learning model based on baseline CT imaging of melanoma lesions had limited value for predicting checkpoint inhibitor treatment outcomes**. Furthermore, this approach was unable to add information over a clinical model. The predictive value of the deep learning model was very comparable to a radiomics model, indicating that the predefined features of a handcrafted radiomics approach are not the limiting factor. Instead, the limited predictive power suggests a lack of predictive information regarding checkpoint inhibitor response in the single-energy CT images of melanoma lesions. Future research may investigate spectral CT imaging, or body composition metrics extracted from baseline CT imaging. Furthermore, research in other modalities remains necessary to move towards accurate baseline predictions of treatment response.

## Data Availability

Due to confidentiality agreements, clinical and imaging data cannot be made available.

## Conflict of interest statement

AvdE has advisory relationships with Bristol-Myers Squibb, MSD Oncology, Amgen, Roche, Novartis, Sanofi, Pfizer, Ipsen, Merck, Pierre Fabre and has received research study grants not related to this paper from Sanofi, Bristol-Myers Squibb, TEVA, Idera and has received travel expenses MSD Oncology, Roche, Pfizer, Sanofi, Pierre Fabre and has received speaker honoraria from BMS and Novartis.

JdG has consultancy/advisory relationships with Bristol Myers Squibb, Pierre Fabre, Servier, MSD, Novartis.

PJ has a research collaboration with Philips Healthcare and Vifor Pharma.

MBS has consultancy/advisory relationships with Pierre Fabre, MSD and Novartis, none related to current work and paid to institute.

EK has consultancy/advisory relationships with Bristol Myers Squibb, Novartis, Merck, Pierre Fabre, Lilly, Bayer, EISAI and Ipsen paid to the institute, and received research grants not related to this paper from Bristol Myers Squibb, Delcath, Novartis and Pierre Fabre.

PD has consultancy/advisory relationships with Paige, Pantarei and Samantree paid to the institution and research grants from Pfizer, none related to current work and paid to institute.

KS has advisory relationships with Bristol Myers Squibb, Novartis, MSD, Pierre Fabre, AbbVie, Sairopsa and received honoraria from Novartis and MSD and research funding from Bristol Myers Squibb, TigaTx and Philips.

TL has received research funding from Philips.

GH consultancy/advisory relationships with Amgen, Bristol-Myers Squibb, Roche, MSD, Pfizer, Novartis, Sanofi, Pierre Fabre and has received research grants from Bristol-Myers Squibb, Seerave. All payments to the Institution.

HW received honoraria from Merck, Astellas, Roche and travel expenses from Ipsen and Astellas

All remaining authors have declared no conflicts of interest.

