## Supplementary files for "Deep learning on CT scans to predict checkpoint inhibitor treatment outcomes in advanced melanoma"

**Supplementary Methods**

**Scaling vs. cropping method for ROI selection**

During preprocessing, selection of a region of interest around the lesions is performed in order to:

1. Improve the signal-to-noise ratio by limiting the view to the lesion and excluding uninformative parts of the CT scan (normal tissue);
2. Provide a uniform input size to the model despite varying lesion sizes and voxel dimensions;
3. Allow for larger batch sizes and therefore less noisy gradients during model training.

Two methods were investigated for selection of the region of interest of the lesion: ‘scaling’ and ‘cropping’. Of these two methods, the ‘cropping’ method was selected based on the results of initial experiments.

For the ‘cropping’ method, the region of interest was selected to be a cube with fixed dimensions, centered on the centroid of the lesion segmentation. These fixed dimensions were either 50mm, 100mm or 150mm. The advantage of this method is that the scale is the same across cases, which means that information about size is preserved. The disadvantage is that the largest lesions do not fully with within the region of interest, which means that information about the edges of these lesions is lost.

For the ‘scaling’ method, the region of interest is obtained in the following way: first, a bounding box of the segmentation is obtained. Subsequently, a margin of 0mm, 10mm or 50mm is added on all sides to this bounding box. For every lesion, this results in a cuboid of varying sizes. Next, the two smallest dimensions are increased symmetrically to match the size of the largest dimension, so that a cube is obtained. Lastly, this cube is resampled to match the input size of the deep learning model. The advantage of this method is that the lesion is guaranteed to fit in the region of interest; the disadvantage is that scale is not preserved.

**Deep learning model recalibration**

As described in the main text, the deep learning model is optimized to predict lesion level outcomes. The resulting lesion level predictions are subsequently aggregated to a patient level prediction by taking the minimum, mean or maximum of the predictions per lesion. This aggregation may result in miscalibration: if the ‘minimum’ is used, this may result in predictions which are too low on average. This is offset by recalibrating the output of the model using the following steps:

1. In the inner cross validation loop, the fitted model is used to make predictions on the validation fold. This validation fold has been used to monitor training and activate early stopping, but is unseen by the model. This results in a prediction for every lesion in the training set.
2. These predictions per lesion are aggregated to a patient level prediction for every patient in the training set using the selected aggregation method (minimum, mean or maximum).
3. An unpenalized logistic regression is fit on the predictions per patient to predict the target outcome (e.g. clinical benefit). This logistic regression counteracts the miscalibration due to aggregating by re-estimating offset and slope.
4. The deep learning ensemble (consisting of five models fitted on the data from the training centers) is used to make predictions for every lesion of the test center. These five predictions per lesion are combined by taking the mean of all predictions.
5. Per lesion predictions in the data from the test center are combined to per-patient predictions using the selected aggregation method (minimum, mean or maximum).
6. The logistic regression fitted on the training data is used to recalibrate these per-patient predictions.

**Clinical model**

The deep learning model was compared to a model built on clinical predictors, consisting of the following variables and corresponding categories:

- **Liver metastases:** present vs. absent vs. missing
- **Brain metastases**: present vs. asymptomatic vs. symptomatic vs. missing
- **ECOG performance status:** 0 vs. 1 vs. 2-4 vs. missing
- **LDH:** normal vs. 1-2x upper limit of normal vs. ≥3x upper limit of normal vs. missing

Missing values were encoded as a separate level. As continuous values were not available for LDH, categories were used instead. A logistic regression with L2-regularization was fitted on these four variables with a total of 15 levels. During the model training phase, the optimal L2 penalty was estimated using a cross validation on the training set. The L2 penalty was subsequently fixed on this optimal value, and the model was retrained on the entire training set. During the inference phase, this trained model was evaluated on an independent test set. This process was repeated for all iterations of the leave-one-center-out cross validation.

**Combination model**

The combination model is based on the output of the deep learning model and the clinical model. The model is built by the following steps:

1. Per patient predictions for patients in the training centers are obtained from the deep learning model by steps 1 and 2 under ‘Deep learning model recalibration’.
2. Using the optimal L2 penalty (determined using the method under ‘Clinical model’), independent predictions from the clinical model are obtained for the training set using a cross validation.
3. The combination model, consisting of an unpenalized logistic regression, is fitted on the per patient predictions of the deep learning model and clinical model.
4. Per patient predictions are obtained from the deep learning model for every patient in the test center by steps 4 and 5 under ‘Deep learning model recalibration’. Similarly, per patient predictions are obtained from the clinical model.
5. Based on these predictions, the combination model outputs a combined prediction.

**Comparison of AUROCs between models**

In the absence of an alternative to the DeLong’s test [1] for comparing AUROCs in a cross validation setup, the following method was used to test for significant differences in discrimination between two models. In every validation fold, the difference in AUROC for the two models was calculated. An estimate of the standard deviation of this difference as obtained by bootstrapping the samples in the fold. The results were pooled across all validation folds using a random effects model. If the 95% confidence interval of the pooled difference did not include 0, differences were said to be significant. It must be noted that this method does not account for the dependence between folds; the resulting confidence intervals may therefore be, on average, too small [2].

**Supplementary Table 1 - Hyperparameters**

| **Hyperparameter** | **Explored values** | **Selected in final model** |
| --- | --- | --- |
| ROI input dimension | - 3D - 2.5D [3] | 3D |
| Lesion to patient level prediction aggregation function | - Minimum - Mean - Maximum | Minimum |
| Optimizer | - Adam - SGD | Adam |
| Weight decay | - 0 - 10^-9^ - 10^-7^ - 10^-5^ - 10^-3^ - 10^-1^ | 10^-7^ |
| Model | - Densenet121 - Densenet169 - Densenet201 - Efficientnet-b0 - Efficientnet-b1 - Efficientnet-b2 - SEResNet50 - SEResNet101 - SEResNet152 - SEResNext50 - SEResNext101 | SEResNet50 |
| Dropout | Uniform distribution from 0 to 0.7 | 0 |
| Momentum (only for SGD optimizer) | - 0 - 0.5 - 0.9 - 0.99 | Disabled |
| Pretraining | - None - ImageNet | None |
| Sampler | - Normal - One patient per batch - Stratified for label - Stratified for organ and label | Normal |
| ROI selection method | - Crop - Scale | Crop |
| ROI size (mm) (only for ‘crop’ method) | - 50 - 100 - 150 | 100 |
| ROI margin (mm) (only for ‘scale’ method) | - 0 - 10 - 50 | None |
| Maximum learning rate | Log-uniform distribution from 10^-6^ to 10^-2^ | 10^-5^ |
| Minimum learning rate | Fixed | 10^-7^ |
| Max epochs | Fixed | 100 |
| Patience for early stopping | Fixed | 10 |
| Period of cosine annealing (epochs) | Fixed | 10 |

**Supplementary Table 2 - Availability of lesion outcomes at 3, 6 and 9 months**

|  | Baseline | 3 months | 6 months | 9 months | Last available followup |
| --- | --- | --- | --- | --- | --- |
| Total lesions, n (%) | 2771 (100%) | 2128 (77%) | 1708 (61%) | 1538 (56%) | 2128 (77%) |
| Has response, n (%) |  | 917 (43%) | 1182 (69%) | 1234 (80%) | 1176 (55%) |
| Has benefit, n (%) |  | 1649 (77%) | 1523 (89%) | 1448 (94%) | 1697 (80%) |
| Missing, n |  | 643 (23%) | 1063 (38%) | 1233 (44%) | 643 (23%) |

**Supplementary Table 3 - Characteristics of in- and excluded patients**

|  |  | Included | Excluded |
| --- | --- | --- | --- |
| n |  | 730 | 617 |
| Age, median [Q1,Q3] |  | 68.0 [58.0,75.0] | 67.0 [57.0,76.0] |
| Sex, n (%) | Female | 285 (39.0) | 230 (37.3) |
|  | Male | 445 (61.0) | 387 (62.7) |
| Therapy, n (%) | Anti-PD1 | 458 (62.7) | 418 (67.7) |
|  | Ipilimumab & Nivolumab | 272 (37.3) | 199 (32.3) |
| Stage, n (%) | IIIC | 28 (3.8) | 82 (13.3) |
|  | IV M1a | 56 (7.7) | 53 (8.6) |
|  | IV M1b | 114 (15.6) | 74 (12.0) |
|  | IV M1c | 344 (47.1) | 253 (41.0) |
|  | IV M1d | 182 (24.9) | 149 (24.1) |
|  | missing | 6 (0.8) | 6 (1.0) |
| ECOG performance status, n (%) | 0 | 356 (48.8) | 276 (44.7) |
|  | 1 | 271 (37.1) | 269 (43.6) |
|  | 2-4 | 73 (10.0) | 41 (6.6) |
|  | missing | 30 (4.1) | 31 (5.0) |
| Brain metastases, n (%) | absent | 497 (68.1) | 360 (58.3) |
|  | asymptomatic | 94 (12.9) | 93 (15.1) |
|  | symptomatic | 88 (12.1) | 56 (9.1) |
|  | missing | 51 (7.0) | 108 (17.5) |
| Liver metastases, n (%) | absent | 471 (64.5) | 400 (64.8) |
|  | present | 224 (30.7) | 123 (19.9) |
|  | missing | 35 (4.8) | 94 (15.2) |
| LDH, n (%) | normal | 459 (62.9) | 423 (68.6) |
|  | 1-2x ULN | 199 (27.3) | 148 (24.0) |
|  | ≥3x ULN | 62 (8.5) | 38 (6.2) |
|  | missing | 10 (1.4) | 8 (1.3) |
| Number of affected organs, n (%) | <3 | 432 (59.2) | 392 (63.5) |
|  | ≥3 | 298 (40.8) | 225 (36.5) |
| Best overall response, n (%) | Complete response | 94 (12.9) | 128 (20.7) |
|  | Partial response | 279 (38.2) | 181 (29.3) |
|  | Stable disease | 115 (15.8) | 103 (16.7) |
|  | Progressive disease | 237 (32.5) | 199 (32.3) |
|  | Death | 5 (0.7) | 6 (1.0) |
| Clinical benefit, n (%) | benefit | 435 (59.6) | 345 (62.6) |
|  | no benefit | 295 (40.4) | 206 (37.4) |
| Objective response, n (%) | response | 373 (51.1) | 305 (55.4) |
|  | no response | 357 (48.9) | 246 (44.6) |
| Abbreviations  ECOG=Eastern Cooperative Oncology Group LDH=lactate dehydrogenase, ULN=upper limit of normal, defined as 250 IU/L | | | |

**Supplementary Table 4 - CT acquisition parameters for included patients**

|  |  | Amphia | Isala Zwolle | LUMC | Maxima MC | MST | Radboudumc | UMCG | UMCU | Amsterdam UMC | Zuyderland |
| --- | --- | --- | --- | --- | --- | --- | --- | --- | --- | --- | --- |
| n |  | 50 | 96 | 72 | 54 | 23 | 82 | 110 | 93 | 119 | 29 |
| Current, median [Q1,Q3] |  | 120.0 [91.0,255.0] | 271.0 [195.0,377.0] | 160.0 [110.0,283.0] | 312.0 [235.0,380.0] | 397.5 [249.2,525.0] | 277.0 [176.0,409.2] | 401.0 [195.0,613.0] | 234.5 [179.5,278.5] | 229.0 [143.8,327.8] | 272.5 [205.2,350.0] |
| Voltage, median [Q1,Q3] |  | 100.0 [100.0,100.0] | 120.0 [120.0,120.0] | 120.0 [120.0,120.0] | 100.0 [100.0,100.0] | 100.0 [100.0,120.0] | 100.0 [100.0,120.0] | 100.0 [100.0,100.0] | 120.0 [120.0,120.0] | 110.0 [100.0,120.0] | 120.0 [120.0,120.0] |
| Slice thickness, median [Q1,Q3] |  | 3.0 [3.0,3.0] | 1.0 [0.9,1.2] | 1.0 [1.0,1.0] | 3.0 [3.0,3.0] | 3.0 [3.0,3.0] | 1.0 [1.0,1.0] | 2.0 [1.0,2.0] | 0.9 [0.9,1.0] | 1.0 [0.9,2.0] | 3.0 [2.0,5.0] |
| Pixel spacing, median [Q1,Q3] |  | 0.8 [0.7,0.9] | 0.8 [0.7,0.9] | 0.8 [0.8,0.8] | 0.5 [0.5,0.7] | 0.8 [0.8,0.8] | 0.8 [0.7,0.9] | 0.8 [0.7,0.8] | 0.8 [0.7,0.8] | 0.8 [0.7,1.0] | 0.8 [0.8,0.9] |
| Vendor, n (%) | GE MEDICAL SYSTEMS | 2 (4.0) | 6 (6.7) | 2 (3.7) | 2 (3.9) | 1 (4.3) | 2 (2.8) | 3 (3.3) |  | 9 (12.0) |  |
|  | Philips | 6 (12.0) | 70 (78.7) | 2 (3.7) | 49 (96.1) |  | 13 (18.3) | 1 (1.1) | 35 (76.1) | 21 (28.0) |  |
|  | Philips Medical Systems | 1 (2.0) |  |  |  |  |  |  |  |  |  |
|  | SIEMENS | 41 (82.0) | 7 (7.9) | 1 (1.9) |  | 22 (95.7) | 16 (22.5) | 83 (92.2) | 10 (21.7) | 42 (56.0) |  |
|  | TOSHIBA |  | 6 (6.7) | 48 (88.9) |  |  | 40 (56.3) | 2 (2.2) | 1 (2.2) | 3 (4.0) |  |
|  | Canon Medical Systems |  |  | 1 (1.9) |  |  |  |  |  |  |  |
|  | Carestream Health |  |  |  |  |  |  | 1 (1.1) |  |  |  |
| Model, n (%) | Biograph128 | 1 (2.0) |  |  |  |  |  | 10 (11.2) |  | 1 (1.3) |  |
|  | Biograph40 | 1 (2.0) | 2 (2.2) |  |  |  | 4 (5.6) | 8 (9.0) | 2 (4.3) | 1 (1.3) |  |
|  | Biograph64 | 21 (42.0) |  |  |  |  |  | 12 (13.5) |  | 8 (10.7) |  |
|  | GEMINI TF TOF 16 | 1 (2.0) |  |  |  |  |  |  |  |  |  |
|  | Ingenuity CT | 1 (2.0) | 16 (18.0) |  | 28 (54.9) |  | 1 (1.4) |  |  |  |  |
|  | Ingenuity TF PET/CT | 2 (4.0) | 7 (7.9) |  |  |  |  |  |  | 5 (6.7) |  |
|  | LightSpeed VCT | 2 (4.0) |  | 1 (1.9) |  |  |  |  |  |  |  |
|  | SOMATOM Definition AS | 14 (28.0) | 1 (1.1) |  |  | 4 (17.4) | 3 (4.2) | 17 (19.1) | 3 (6.5) | 2 (2.7) |  |
|  | SOMATOM Definition Flash | 2 (4.0) | 1 (1.1) | 1 (1.9) |  | 14 (60.9) | 3 (4.2) | 9 (10.1) | 2 (4.3) | 1 (1.3) |  |
|  | SOMATOM Edge Plus | 2 (4.0) |  |  |  |  |  |  |  |  |  |
|  | TruFlight Select | 1 (2.0) |  |  |  |  |  |  |  |  |  |
|  | iCT 256 | 2 (4.0) | 15 (16.9) | 1 (1.9) | 20 (39.2) |  | 7 (9.9) |  | 17 (37.0) | 10 (13.3) |  |
|  | Aquilion ONE |  | 1 (1.1) | 30 (55.6) |  |  | 17 (23.9) |  |  | 1 (1.3) |  |
|  | Aquilion PRIME |  | 5 (5.6) |  |  |  |  |  |  |  |  |
|  | Brilliance 16 |  | 1 (1.1) |  |  |  |  | 1 (1.1) |  |  |  |
|  | Discovery STE |  | 4 (4.5) |  |  |  |  |  |  |  |  |
|  | Ingenuity TF PET/CT |  | 23 (25.8) |  |  |  |  |  |  |  |  |
|  | Optima CT660 |  | 2 (2.2) |  |  |  | 2 (2.8) | 3 (3.4) |  |  |  |
|  | SOMATOM Definition Edge |  | 3 (3.4) |  |  |  | 1 (1.4) |  |  |  |  |
|  | Vereos PET/CT |  | 8 (9.0) |  |  |  |  |  |  | 3 (4.0) |  |
|  | Aquilion |  |  | 19 (35.2) |  |  | 6 (8.5) | 2 (2.2) | 1 (2.2) | 2 (2.7) |  |
|  | Discovery MI |  |  | 1 (1.9) |  | 1 (4.3) |  |  |  |  |  |
|  | GEMINI TF TOF 64T |  |  | 1 (1.9) |  |  |  |  |  |  |  |
|  | Discovery 710 |  |  |  | 2 (3.9) |  |  |  |  |  |  |
|  | Mx8000 IDT 16 |  |  |  | 1 (2.0) |  |  |  |  |  |  |
|  | Biograph 40 |  |  |  |  | 2 (8.7) | 1 (1.4) | 13 (14.6) |  | 1 (1.3) |  |
|  | SOMATOM Definition AS+ |  |  |  |  | 2 (8.7) |  |  |  | 1 (1.3) |  |
|  | Aquilion Precision |  |  |  |  |  | 17 (23.9) |  |  |  |  |
|  | Brilliance 40 |  |  |  |  |  | 2 (2.8) |  |  |  |  |
|  | Brilliance 64 |  |  |  |  |  | 2 (2.8) |  | 6 (13.0) | 2 (2.7) |  |
|  | GEMINI TF TOF 64 |  |  |  |  |  | 1 (1.4) |  |  |  |  |
|  | Sensation 64 |  |  |  |  |  | 3 (4.2) |  |  | 1 (1.3) |  |
|  | syngo.via.VB20A |  |  |  |  |  | 1 (1.4) |  |  |  |  |
|  | Biograph 64 |  |  |  |  |  |  | 6 (6.7) |  |  |  |
|  | Biograph20 |  |  |  |  |  |  | 2 (2.2) |  |  |  |
|  | Biograph40_mCT 4R |  |  |  |  |  |  | 1 (1.1) |  |  |  |
|  | SOMATOM Force |  |  |  |  |  |  | 4 (4.5) | 2 (4.3) | 9 (12.0) |  |
|  | SOMATOM go.All |  |  |  |  |  |  | 1 (1.1) |  |  |  |
|  | Emotion 6 |  |  |  |  |  |  |  | 1 (2.2) |  |  |
|  | IQon - Spectral CT |  |  |  |  |  |  |  | 12 (26.1) |  |  |
|  | Biograph 16 |  |  |  |  |  |  |  |  | 9 (12.0) |  |
|  | Biograph128Edge |  |  |  |  |  |  |  |  | 3 (4.0) |  |
|  | Discovery CT750 HD |  |  |  |  |  |  |  |  | 9 (12.0) |  |
|  | Ingenuity Core 128 |  |  |  |  |  |  |  |  | 1 (1.3) |  |
|  | SOMATOM Drive |  |  |  |  |  |  |  |  | 5 (6.7) |  |

**Supplementary Table 5 - Patient characteristics per treatment subgroup**

|  |  | Anti-PD1 | Ipilimumab & Nivolumab |
| --- | --- | --- | --- |
| n |  | 458 | 272 |
| Age, median [Q1,Q3] |  | 69.0 [61.0,76.8] | 63.0 [53.0,72.0] |
| Sex, n (%) | Female | 180 (39.3) | 105 (38.6) |
|  | Male | 278 (60.7) | 167 (61.4) |
| Stage, n (%) | IIIC | 25 (5.5) | 3 (1.1) |
|  | IV M1a | 52 (11.4) | 4 (1.5) |
|  | IV M1b | 99 (21.6) | 15 (5.5) |
|  | IV M1c | 208 (45.4) | 136 (50.0) |
|  | IV M1d | 70 (15.3) | 112 (41.2) |
|  | missing | 4 (0.9) | 2 (0.7) |
| ECOG performance status, n (%) | 0 | 244 (53.3) | 112 (41.2) |
|  | 1 | 158 (34.5) | 113 (41.5) |
|  | 2-4 | 37 (8.1) | 36 (13.2) |
|  | missing | 19 (4.1) | 11 (4.0) |
| Brain metastases, n (%) | absent | 350 (76.4) | 147 (54.0) |
|  | asymptomatic | 38 (8.3) | 56 (20.6) |
|  | symptomatic | 32 (7.0) | 56 (20.6) |
|  | missing | 38 (8.3) | 13 (4.8) |
| Liver metastases, n (%) | absent | 327 (71.4) | 144 (52.9) |
|  | present | 101 (22.1) | 123 (45.2) |
|  | missing | 30 (6.6) | 5 (1.8) |
| LDH, n (%) | normal | 349 (76.2) | 110 (40.4) |
|  | 1-2x ULN | 92 (20.1) | 107 (39.3) |
|  | ≥3x ULN | 11 (2.4) | 51 (18.8) |
|  | missing | 6 (1.3) | 4 (1.5) |
| Number of affected organs, n (%) | <3 | 306 (66.8) | 126 (46.3) |
|  | ≥3 | 152 (33.2) | 146 (53.7) |
| Best overall response, n (%) | Complete response | 77 (16.8) | 17 (6.2) |
|  | Partial response | 164 (35.8) | 115 (42.3) |
|  | Stable disease | 80 (17.5) | 35 (12.9) |
|  | Progressive disease | 133 (29.0) | 104 (38.2) |
|  | Death | 4 (0.9) | 1 (0.4) |
| Clinical benefit, n (%) | benefit | 282 (61.6) | 153 (56.2) |
|  | no benefit | 176 (38.4) | 119 (43.8) |
| Objective response, n (%) | response | 241 (52.6) | 132 (48.5) |
|  | no response | 217 (47.4) | 140 (51.5) |
| Abbreviations  ECOG=Eastern Cooperative Oncology Group LDH=lactate dehydrogenase, ULN=upper limit of normal, defined as 250 IU/L | | | |

**Supplementary Table 6 – Patient characteristics per center**

|  |  | Amphia | Isala Zwolle | LUMC | Maxima MC | MST | Radboudumc | UMCG | UMCU | Amsterdam UMC | Zuyderland MC |
| --- | --- | --- | --- | --- | --- | --- | --- | --- | --- | --- | --- |
| n |  | 50 | 96 | 72 | 54 | 23 | 82 | 110 | 93 | 119 | 29 |
| Age, median [Q1,Q3] |  | 65.0 [54.5,71.0] | 69.0 [59.0,76.0] | 71.0 [62.0,77.2] | 66.0 [58.5,75.0] | 63.0 [53.5,69.5] | 63.0 [54.0,71.0] | 68.0 [57.5,74.8] | 69.0 [58.0,74.0] | 70.0 [58.5,76.0] | 66.0 [55.0,71.0] |
| Sex, n (%) | Female | 25 (50.0) | 34 (35.4) | 23 (31.9) | 19 (35.2) | 6 (26.1) | 31 (37.8) | 47 (42.7) | 33 (35.5) | 53 (44.5) | 12 (41.4) |
|  | Male | 25 (50.0) | 62 (64.6) | 49 (68.1) | 35 (64.8) | 17 (73.9) | 51 (62.2) | 63 (57.3) | 60 (64.5) | 66 (55.5) | 17 (58.6) |
| Stage, n (%) | IIIC | 1 (2.0) | 5 (5.2) | 1 (1.4) | 2 (3.7) | 2 (8.7) | 1 (1.2) | 3 (2.7) | 6 (6.5) | 7 (5.9) |  |
|  | M1a | 7 (14.0) | 10 (10.4) | 5 (6.9) | 6 (11.1) | 2 (8.7) | 7 (8.5) | 7 (6.4) | 2 (2.2) | 5 (4.2) | 5 (17.2) |
|  | M1b | 7 (14.0) | 18 (18.8) | 9 (12.5) | 7 (13.0) | 4 (17.4) | 15 (18.3) | 20 (18.2) | 15 (16.1) | 14 (11.8) | 4 (13.8) |
|  | M1c | 23 (46.0) | 41 (42.7) | 34 (47.2) | 23 (42.6) | 9 (39.1) | 40 (48.8) | 48 (43.6) | 38 (40.9) | 71 (59.7) | 16 (55.2) |
|  | M1d | 12 (24.0) | 22 (22.9) | 22 (30.6) | 15 (27.8) | 6 (26.1) | 18 (22.0) | 30 (27.3) | 32 (34.4) | 21 (17.6) | 4 (13.8) |
|  | missing |  |  | 1 (1.4) | 1 (1.9) |  | 1 (1.2) | 2 (1.8) |  | 1 (0.8) |  |
| ECOG performance status, n (%) | 0 | 27 (54.0) | 68 (70.8) | 27 (37.5) | 44 (81.5) | 5 (21.7) | 22 (26.8) | 68 (61.8) | 21 (22.6) | 62 (52.1) | 12 (41.4) |
|  | 1 | 19 (38.0) | 15 (15.6) | 35 (48.6) | 4 (7.4) | 12 (52.2) | 54 (65.9) | 25 (22.7) | 45 (48.4) | 47 (39.5) | 14 (48.3) |
|  | 2-4 | 1 (2.0) | 11 (11.5) | 3 (4.2) | 4 (7.4) | 4 (17.4) | 6 (7.3) | 13 (11.8) | 22 (23.7) | 9 (7.6) |  |
|  | missing | 3 (6.0) | 2 (2.1) | 7 (9.7) | 2 (3.7) | 2 (8.7) |  | 4 (3.6) | 5 (5.4) | 1 (0.8) | 3 (10.3) |
| Brain metastases, n (%) | absent | 36 (72.0) | 67 (69.8) | 46 (63.9) | 28 (51.9) | 15 (65.2) | 61 (74.4) | 74 (67.3) | 55 (59.1) | 88 (73.9) | 25 (86.2) |
|  | asymptomatic | 9 (18.0) | 11 (11.5) | 10 (13.9) | 4 (7.4) | 4 (17.4) | 10 (12.2) | 18 (16.4) | 14 (15.1) | 12 (10.1) | 2 (6.9) |
|  | symptomatic | 3 (6.0) | 11 (11.5) | 12 (16.7) | 11 (20.4) | 2 (8.7) | 8 (9.8) | 12 (10.9) | 18 (19.4) | 9 (7.6) | 2 (6.9) |
|  | missing | 2 (4.0) | 7 (7.3) | 4 (5.6) | 11 (20.4) | 2 (8.7) | 3 (3.7) | 6 (5.5) | 6 (6.5) | 10 (8.4) |  |
| Liver metastases, n (%) | absent | 38 (76.0) | 63 (65.6) | 51 (70.8) | 34 (63.0) | 14 (60.9) | 53 (64.6) | 73 (66.4) | 64 (68.8) | 59 (49.6) | 20 (69.0) |
|  | missing | 1 (2.0) | 5 (5.2) | 3 (4.2) | 2 (3.7) | 3 (13.0) | 1 (1.2) | 5 (4.5) | 6 (6.5) | 9 (7.6) |  |
|  | present | 11 (22.0) | 28 (29.2) | 18 (25.0) | 18 (33.3) | 6 (26.1) | 28 (34.1) | 32 (29.1) | 23 (24.7) | 51 (42.9) | 9 (31.0) |
| LDH, n (%) | normal | 32 (64.0) | 59 (61.5) | 43 (59.7) | 31 (57.4) | 17 (73.9) | 53 (64.6) | 78 (70.9) | 54 (58.1) | 73 (61.3) | 17 (58.6) |
|  | 1-2x ULN | 17 (34.0) | 26 (27.1) | 23 (31.9) | 16 (29.6) | 3 (13.0) | 21 (25.6) | 22 (20.0) | 33 (35.5) | 31 (26.1) | 7 (24.1) |
|  | ≥3x ULN | 1 (2.0) | 8 (8.3) | 5 (6.9) | 6 (11.1) | 3 (13.0) | 8 (9.8) | 9 (8.2) | 6 (6.5) | 11 (9.2) | 5 (17.2) |
|  | missing |  | 3 (3.1) | 1 (1.4) | 1 (1.9) |  |  | 1 (0.9) |  | 4 (3.4) |  |
| Number of affected organs, n (%) | <3 | 31 (62.0) | 67 (69.8) | 38 (52.8) | 31 (57.4) | 14 (60.9) | 51 (62.2) | 60 (54.5) | 59 (63.4) | 62 (52.1) | 17 (58.6) |
|  | ≥3 | 19 (38.0) | 29 (30.2) | 34 (47.2) | 23 (42.6) | 9 (39.1) | 31 (37.8) | 50 (45.5) | 34 (36.6) | 57 (47.9) | 12 (41.4) |
| Best overall response, n (%) | Complete response | 4 (8.0) | 11 (11.5) | 10 (13.9) | 4 (7.4) | 3 (13.0) | 13 (15.9) | 21 (19.1) | 13 (14.0) | 9 (7.6) | 4 (13.8) |
|  | Partial response | 24 (48.0) | 36 (37.5) | 24 (33.3) | 25 (46.3) | 10 (43.5) | 31 (37.8) | 34 (30.9) | 30 (32.3) | 55 (46.2) | 10 (34.5) |
|  | Stable disease | 10 (20.0) | 15 (15.6) | 6 (8.3) | 3 (5.6) | 2 (8.7) | 11 (13.4) | 28 (25.5) | 18 (19.4) | 16 (13.4) | 6 (20.7) |
|  | Progressive disease | 12 (24.0) | 34 (35.4) | 32 (44.4) | 21 (38.9) | 8 (34.8) | 27 (32.9) | 25 (22.7) | 30 (32.3) | 39 (32.8) | 9 (31.0) |
|  | Death |  |  |  | 1 (1.9) |  |  | 2 (1.8) | 2 (2.2) |  |  |
| Clinical benefit, n (%) | benefit | 31 (62.0) | 54 (56.2) | 36 (50.0) | 31 (57.4) | 12 (52.2) | 48 (58.5) | 67 (60.9) | 53 (57.0) | 69 (58.0) | 17 (58.6) |
|  | no benefit | 19 (38.0) | 42 (43.8) | 36 (50.0) | 23 (42.6) | 11 (47.8) | 34 (41.5) | 43 (39.1) | 40 (43.0) | 50 (42.0) | 12 (41.4) |
| Objective response, n (%) | response | 28 (56.0) | 47 (49.0) | 34 (47.2) | 29 (53.7) | 13 (56.5) | 44 (53.7) | 55 (50.0) | 43 (46.2) | 64 (53.8) | 14 (48.3) |
|  | no response | 22 (44.0) | 49 (51.0) | 38 (52.8) | 25 (46.3) | 10 (43.5) | 38 (46.3) | 55 (50.0) | 50 (53.8) | 55 (46.2) | 15 (51.7) |

**Supplementary Table 7 - Results of preliminary experiments**

| **Iteration** | **Parameters explored** | **Parameters fixed based on this run** | **Online report** |
| --- | --- | --- | --- |
| #1 | - Aggregation method - ROI selection method - ROI size - ROI margin - Model - Optimizer - Maximum learning rate - Dropout - Weight decay - Pretraining | - Set aggregation method to ‘minimum’ - Set ROI selection method to ‘crop’ | <https://api.wandb.ai/links/premium/4jwux984> |
| #2 | - Momentum - ROI size - Model - Optimizer - Maximum learning rate - Dropout - Weight decay - Pretraining | - Set optimizer to ‘Adam’ | <https://api.wandb.ai/links/premium/vptvwt9s> |
| #3 | - ROI size - Model - Maximum learning rate - Dropout - Weight decay - Pretraining | - Set pretraining to ‘False’ | <https://api.wandb.ai/links/premium/hlw06n7c> |
| #4 | - ROI selection method - ROI margin - ROI size - Model - Maximum learning rate - Dropout - Weight decay | - Excluded options ‘EfficientNet-b0’, ‘EfficientNet-b1’ and ‘EfficientNet-b2’ for model | <https://api.wandb.ai/links/premium/ml1i0qvc> |
| #5 | - Model - ROI size - Dropout - Weight decay | - Excluded options ‘DenseNet121’, ‘DenseNet169’ and ‘DenseNet201’ for model | <https://api.wandb.ai/links/premium/sh4i8a8b> |
| #6 | - Sampler - Model - ROI size - Maximum learning rate - Dropout - Weight decay | - Excluded options ‘SEResNext50’ and ‘SEResNext101’ for model | <https://api.wandb.ai/links/premium/60gk3k3i> |
| #7 | - Dimension - Model - ROI size - Maximum learning rate - Dropout - Weight decay | - Set dimension to ‘3D’ | <https://api.wandb.ai/links/premium/fdbiv2qm> |
| #8 | - Sampler - Model - ROI size - Maximum learning rate - Dropout - Weight decay | - Set sampler to ‘normal’ - Set model to SEResNet50 - Set ROI size to 50 - Set dropout to 0 - Set maximum learning rate to 10^-5^ | <https://api.wandb.ai/links/premium/uunxgvm2> |
| #9 | - Standard deviation of Gaussian noise for augmentation - Weight decay | - Set weight decay to 10^-7^ - Set standard deviation of Gaussian noise for augmentation to 10^-3^ | <https://api.wandb.ai/links/premium/uxwodat0> |

**Supplementary Figure 1 - Difference in AUROC between deep learning and clinical model**

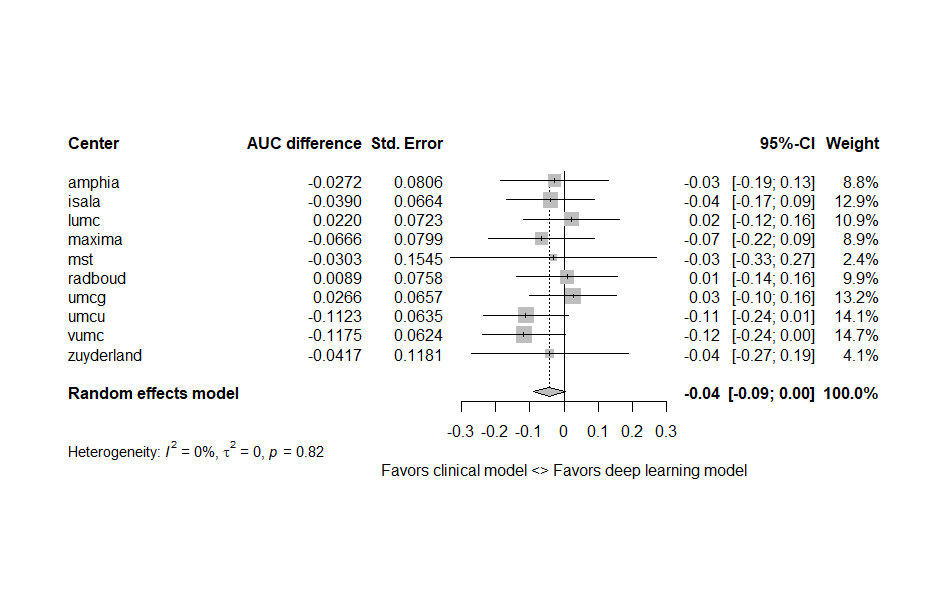

**Supplementary Figure 2 - Difference in AUROC between deep learning and combination model**

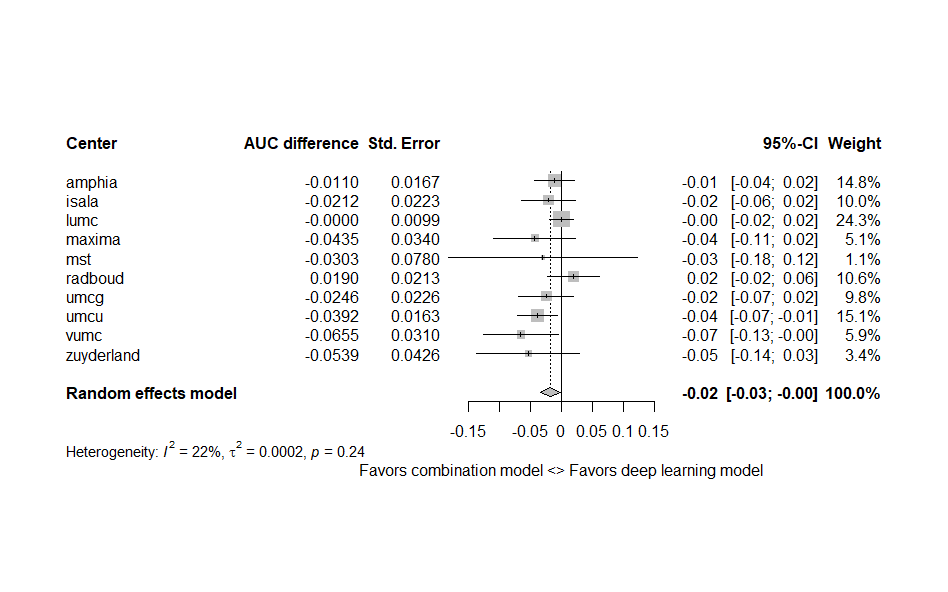

**Supplementary Figure 3 - Difference in AUROC between clinical and combination model**

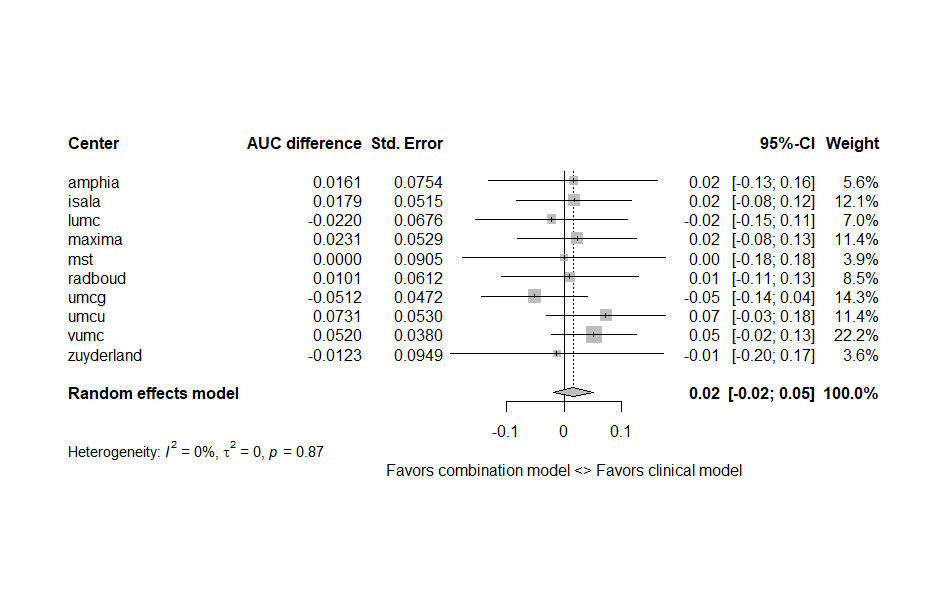

**Supplementary Figure 4 – Receiver operator characteristics curve for predicting response**

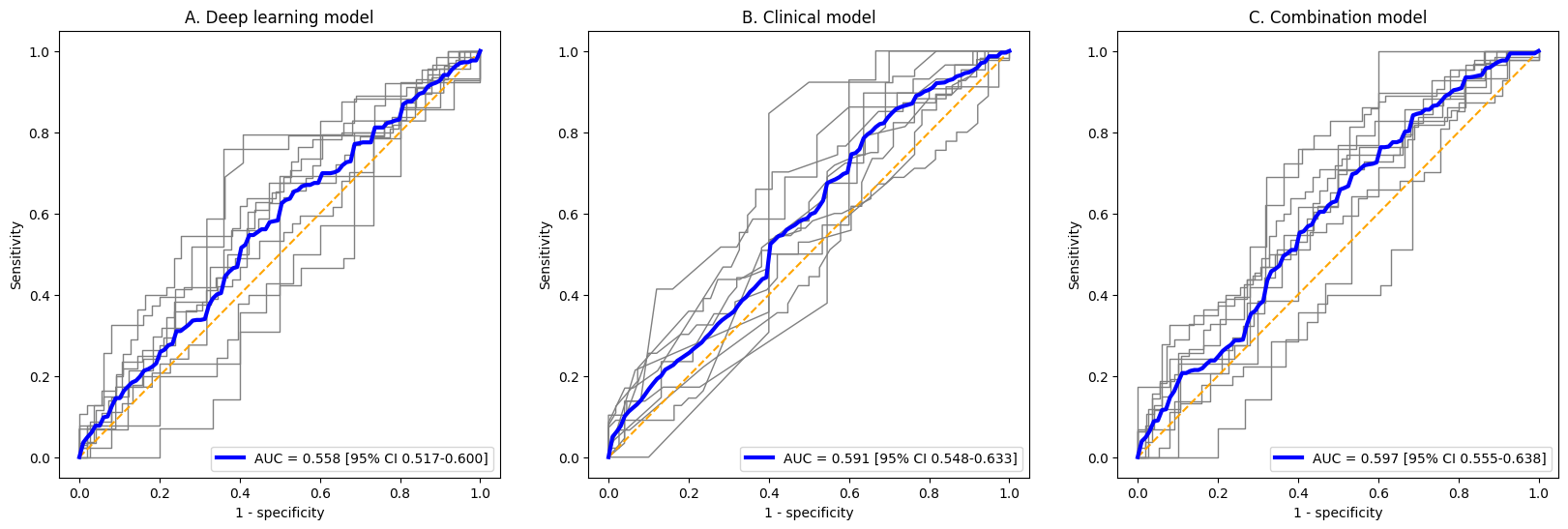

**Supplementary Figure 5 – Calibration curve for predicting response**

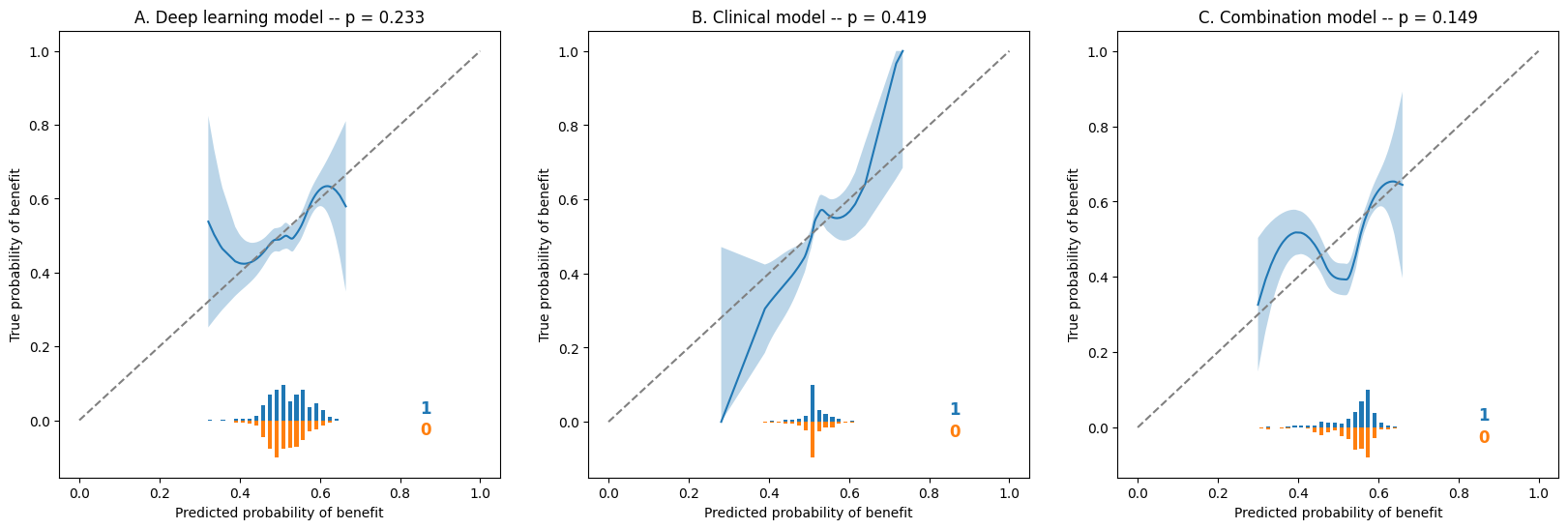

**Supplementary Figure 6 – Receiver operator characteristics curve for predicting clinical benefit in subgroup of patients treated with anti-PD1**

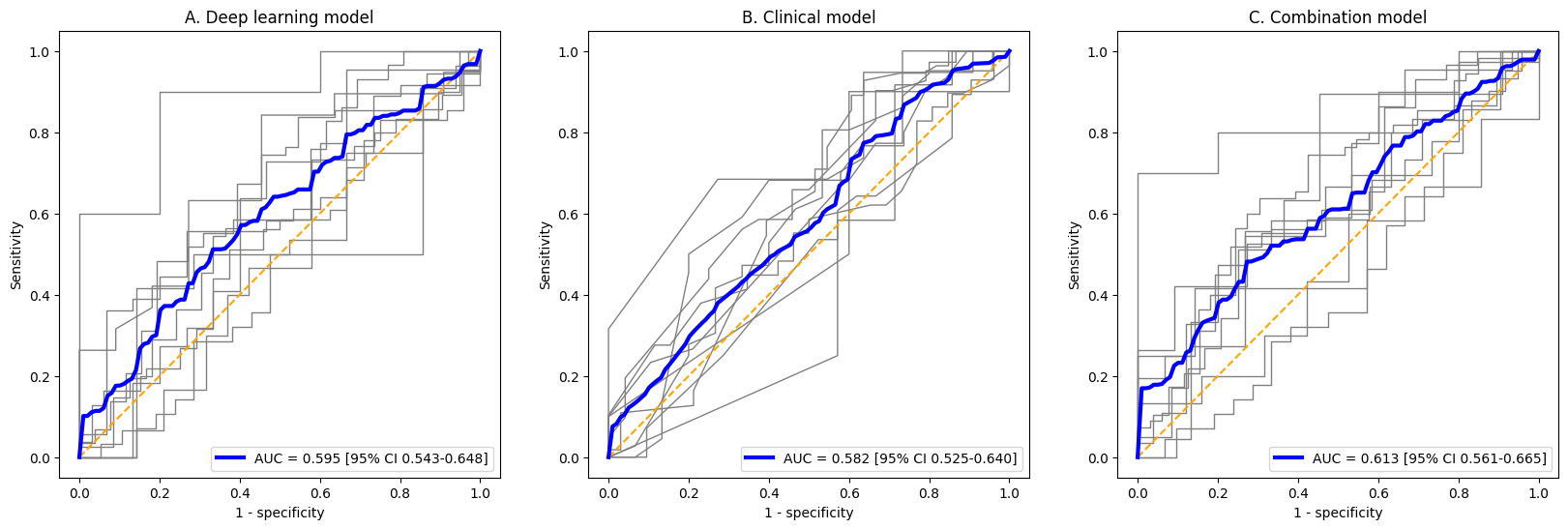

**Supplementary Figure 7 – Calibration curve for predicting clinical benefit in subgroup of patients treated with anti-PD1**

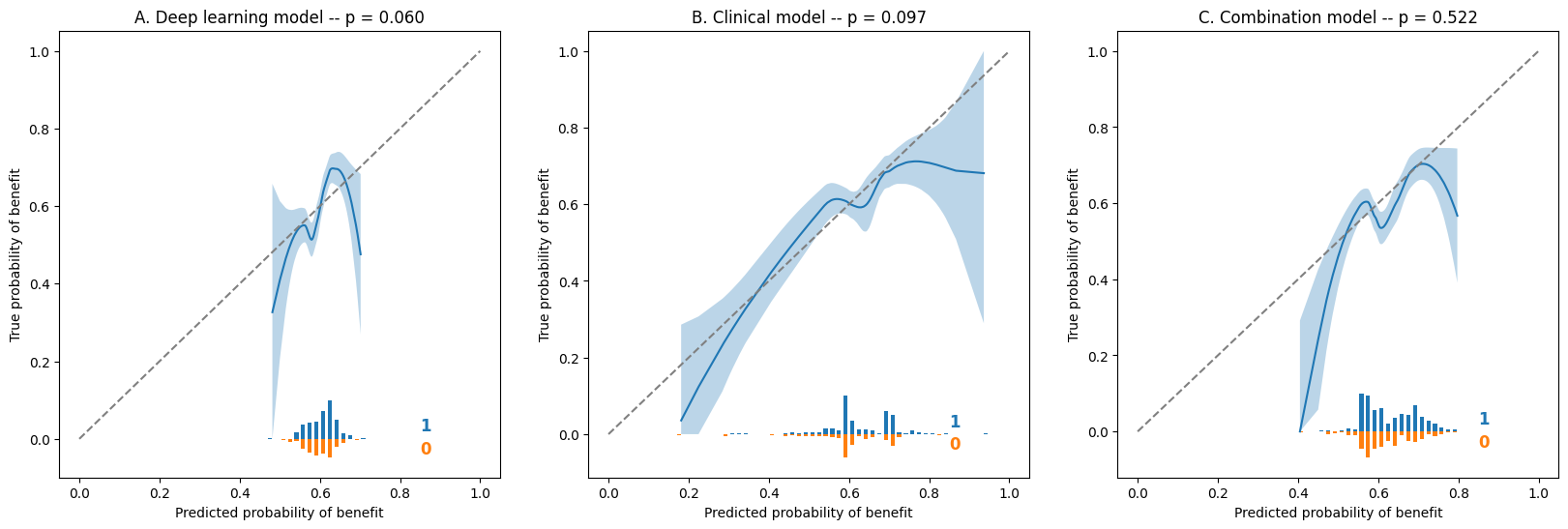

**Supplementary Figure 8 – Receiver operator characteristics curve for predicting clinical benefit in subgroup of patients treated with combination therapy**

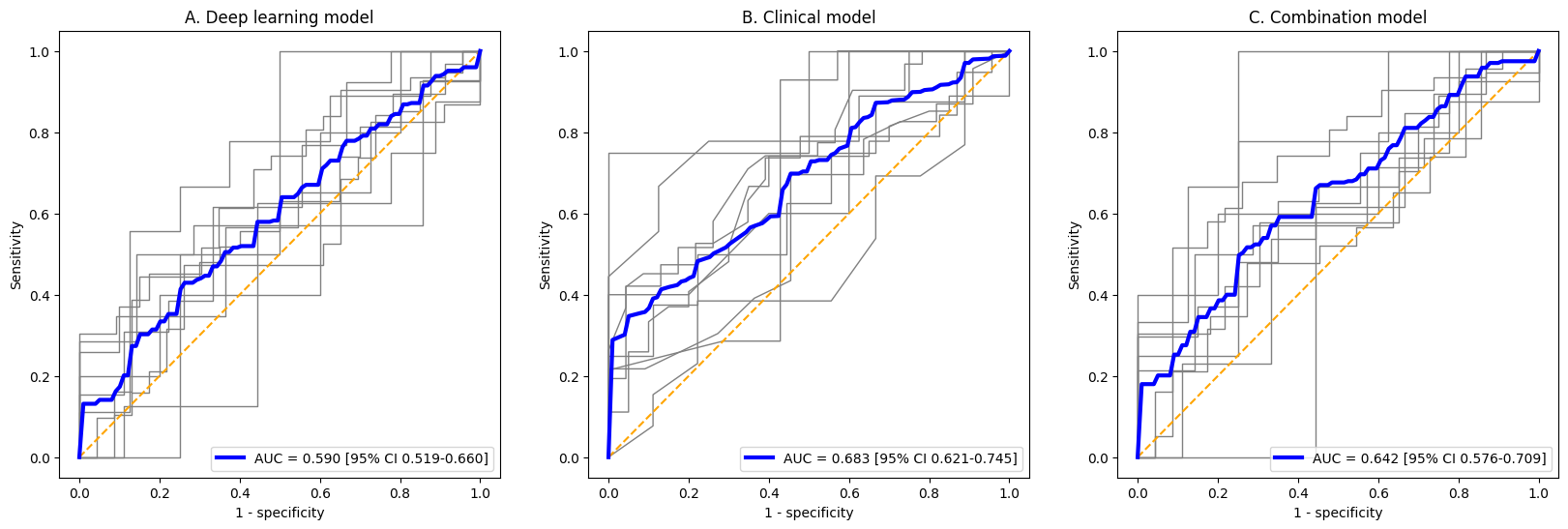

**Supplementary Figure 9 – Calibration curve for predicting clinical benefit in subgroup of patients treated with combination therapy**

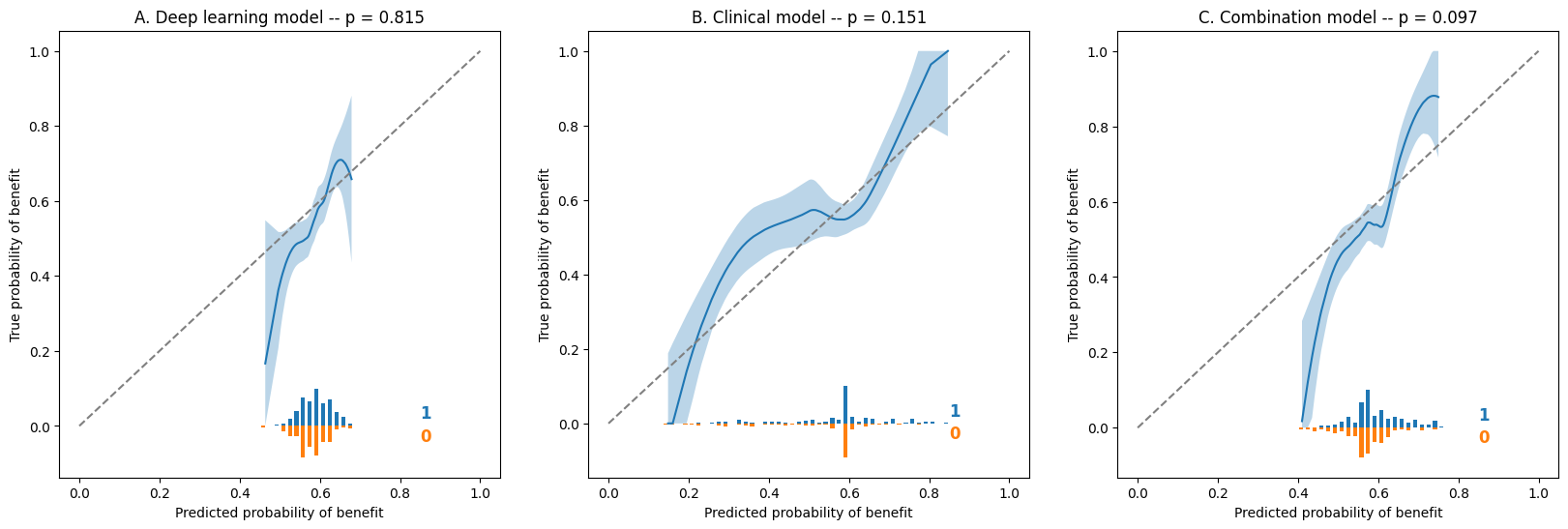

**Supplementary Figure 10 – t-SNE analysis on lesion level of the representation learned by the deep-learning model for predicting clinical benefit (outer fold UMCU, inner fold 0)**

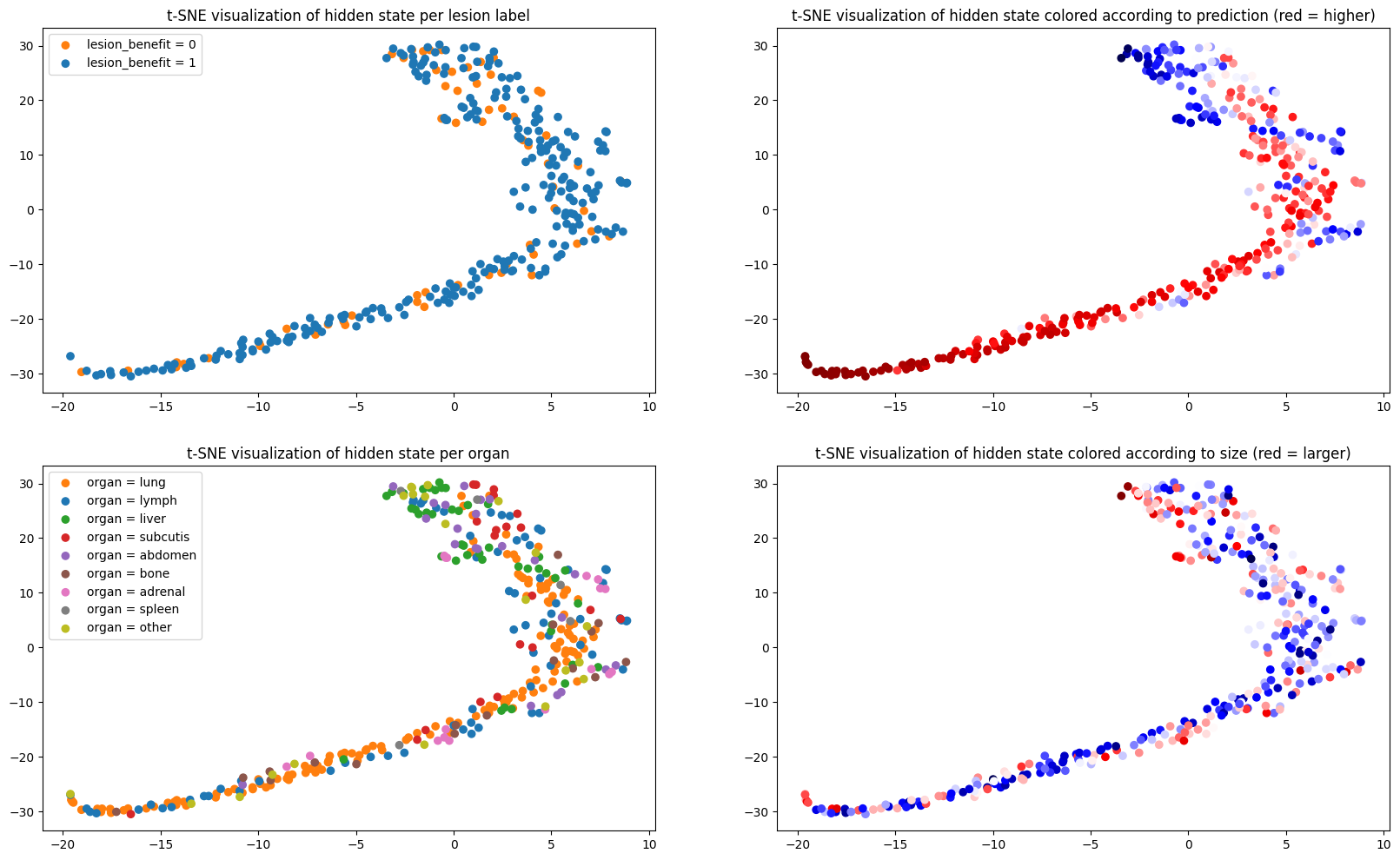

**Supplementary Figure 11 – t-SNE analysis on lesion level of the representation learned by the deep-learning model for predicting clinical benefit (outer fold UMCG, inner fold 0)**

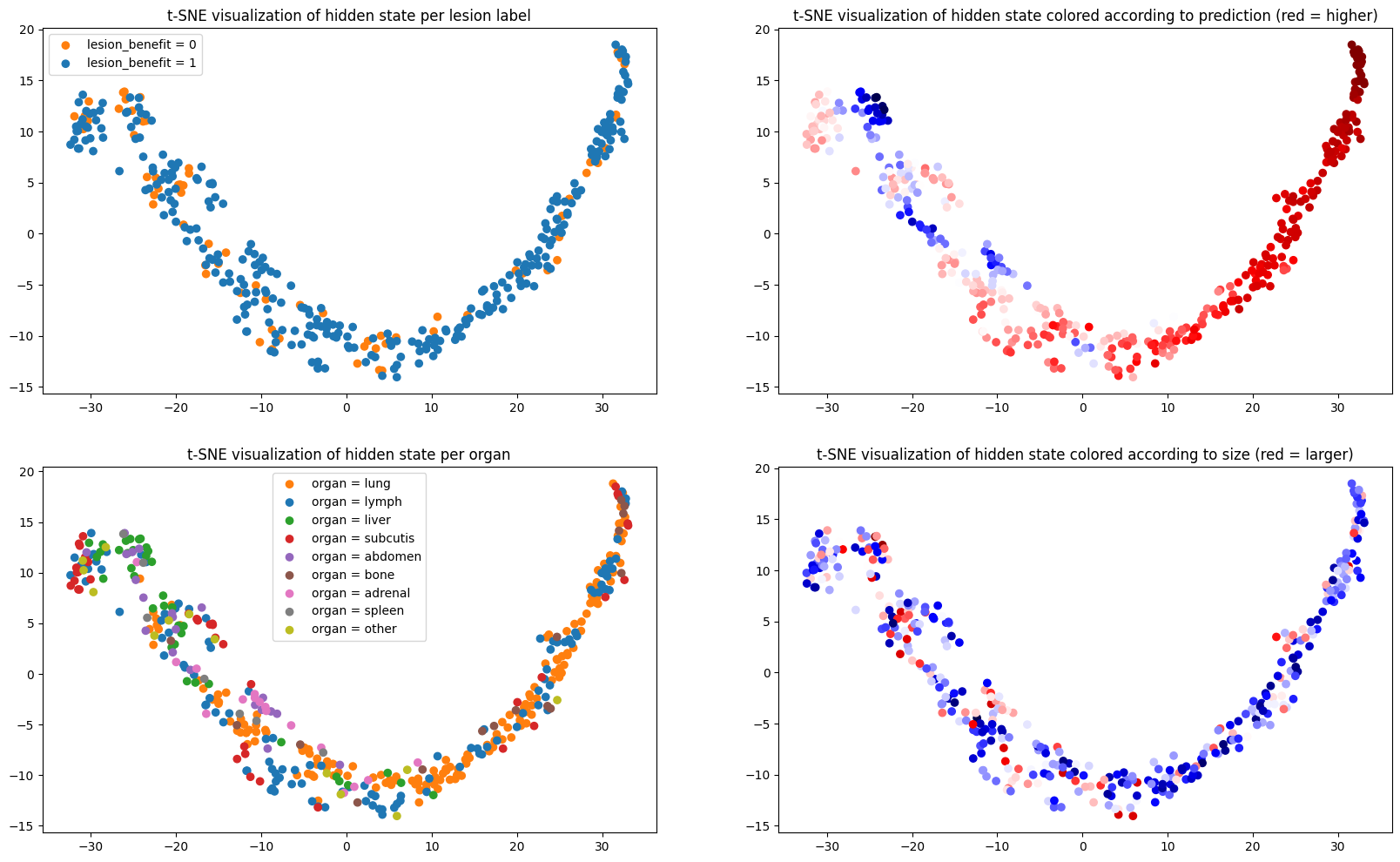
